# An agent-based model to simulate the transmission dynamics of bloodborne pathogens within hospitals

**DOI:** 10.1101/2023.11.14.23298506

**Authors:** Paul Henriot, Mohammed El Kassas, Wagida Anwar, Samia Abdo, Kévin Jean, Laura Temime

## Abstract

Bloodborne pathogens are a major public health concern as they can lead to a variety of medical conditions, including cirrhosis and cancers with significant mortality and morbidity. Three viruses are of major concern: HCV, HBV and HIV. Their transmission is mostly community-associated but the iatrogenic risk of infection is not negligible, even today. Mathematical models are widely used to describe and assess pathogens transmission, within communities and hospitals. Nevertheless, few are focusing on the transmission of pathogens through blood and even fewer on their transmission within hospital as they usually study the risk of community-associated infection in vulnerable populations such as MSM or drug users. Herein, we propose an agent-based SEI (Susceptible-Exposed-Infected) model to explore the transmission dynamics of bloodborne pathogens within hospitals. This model simulates the dynamics of patients between hospital wards, from their admission to discharge, as well as the dynamics of the devices used during at-risk invasive procedures, considering that patient contamination occurs after exposure to a contaminated device. Multiple parameters of the model, such as HCV prevalence, transition probabilities between wards or ward-specific probabilities of undergoing different invasive procedures, were informed with data collected in the University Hospital of Ain Shams in Cairo, Egypt in 2017. We explored the effect of device shortage as well as the effect of random and systematic screening with associated modification in disinfection practices on the risk of infection for patients. By modifying some parameters of the model, we then explored the case of HBV in Ethiopia. In the future, this model could be used to assess the risk of transmission of other bloodborne pathogens in other contexts.

## 1. Introduction

Bloodborne pathogens are a major public health concern as they can lead to a variety of medical conditions, including cirrhosis and cancers with significant mortality and morbidity (Pirozzolo and LeMay, 2007). Three viruses are of major concern: hepatitis B and C viruses (HBV and HCV), and human immunodeficiency virus (HIV). Transmission is mostly community-associated, especially due to unsafe injections during drug use. Nevertheless, iatrogenic transmission may occur during invasive procedures when compliance to infection control measures is imperfect. Recent studies have highlighted the persisting increased risk of HBV and HCV infection in individuals exposed to invasive procedures, with high risk levels in some low-or middle-income countries (2,3).

Mathematical models are widely used to better understand pathogen transmission and assess control strategies, in particular within healthcare settings (4). Nevertheless, models investigating the bloodborne transmission of pathogens remain rare and often focused on community-level epidemics, notably among drug users (5). To our knowledge, no dynamic model has been proposed to specifically explore within-hospital transmission routes of bloodborne viruses and evaluate control measures.

Herein, we present a flexible agent-based model describing movements (from admission to discharge) of patients between wards in a hospital, their exposure to pathogen-contaminated blood through medical devices during invasive procedures, and the ensuing epidemic process. We apply this model to simulate HCV nosocomial transmission, relying on detailed observational data collected in an Egyptian hospital, and extend it to the case of HBV. We explore varying levels of infection control within the hospital, and simulate interventions in order to assess their effect on the risk of infection for hospitalized patients.

## 2. Methods

We developed a stochastic, discrete-time, individual-based model that describes the dynamics of patients within a hospital, from admission to discharge, as well as the dynamics of medical and surgical devices used during at-risk invasive procedures. The model further simulates the between-patient spreading dynamics of a bloodborne pathogen through contaminated devices (Fig 1). Transmission occurs during invasive procedures: device contamination from an infectious patient to an uncontaminated device and patient infection from a contaminated device to a susceptible patient. Herein we describe the mathematical framework of the model as well as its specificities from a user point of view, its parametrization and operation.

**Figure 1.**
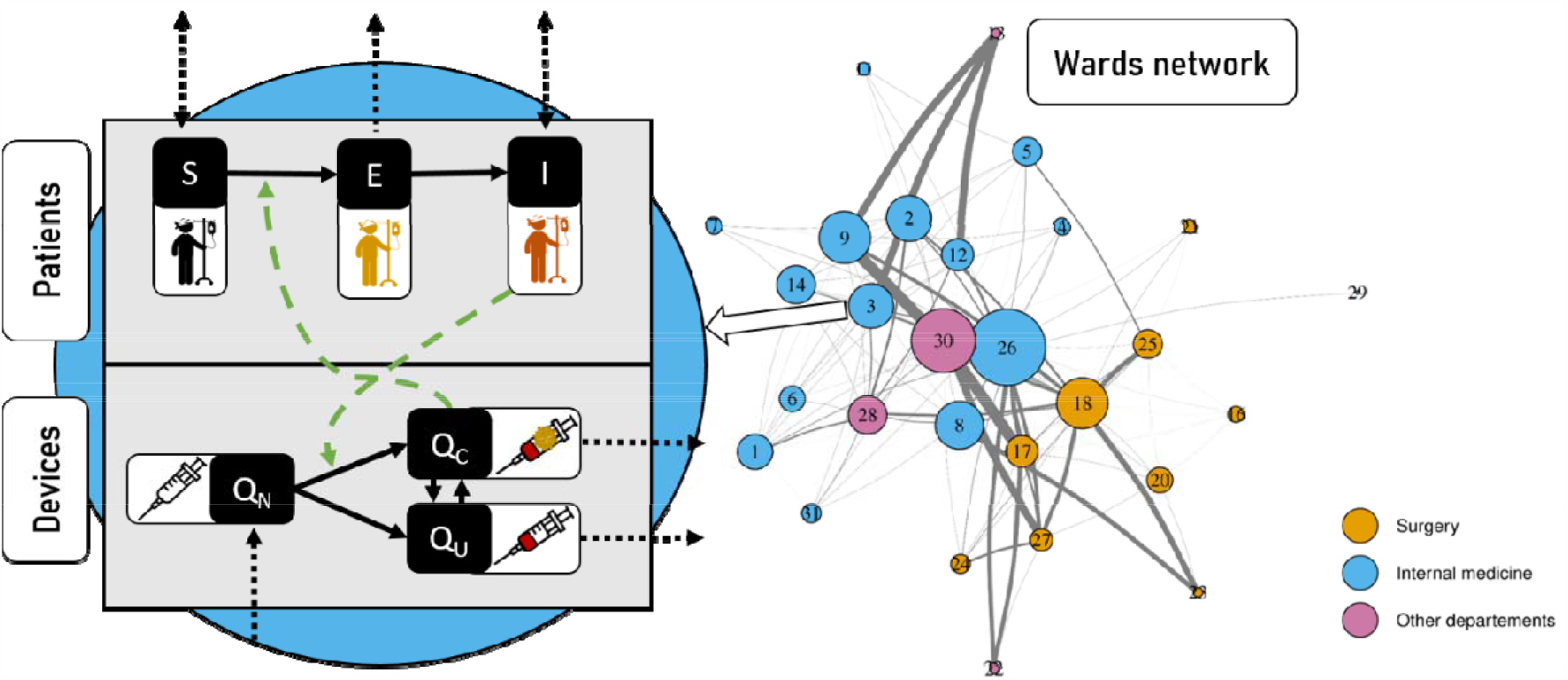
Model framework. In each ward, patients are categorized as either Susceptible (S), Exposed (E) or Infectious (I), while the available devices may either be new (Q_N_) and sterile, previously used but uncontaminated (Q_U_) or previously used and contaminated (Q_C_). The ward network represents the structure of the Ain Shams hospital into 28wards, used as an example for our model application. Each number is associated with a ward, with the numbered list of wards provided in table S1.

### 2.1 Population Dynamics

Table 1A and 1B summarise model parameters. The total hospitalized population is assumed to remain constant over time, at hospital capacity *N*_*pat*_, so that the number of patients leaving the hospital at time *t* is equal to the number of patients entering the hospital at time *t+1*. Patients may belong to any of *N*_*g*_ distinct profiles such as age, place of admission, etc… For each profile *g* and each ward *w* the upon-admission prevalence is defined as 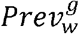.

**Table 1A.** Hospital-associated parameters.

| Entry parameters | Notation | Value |  | Reference |
| --- | --- | --- | --- | --- |
| Initial quantity of new devices $k$ in ward $w$ | $q_{n,k,w}^{init}$ | Tables S7-S10 | | Data |
| Initial quantity of previously used uncontaminated devices $k$ in ward $w$ | $q_{u,k,w}^{init}$ | | | |
| Initial quantity of previously used contaminated devices $k$ in ward $w$ | $q_{c,k,w}^{init}$ | | | |
| Probability of successful sterilization for device of type $k$ | $P_{ster,k}$ | 0.95 (for all devices in the high-resource setting)<br>0.80 (for all devices in the low-resource setting) | | Expert opinion |
| Resupply frequency for device of type $k$ | $f_k$ | Text S1 | | Estimated from data |
|  |  | **Surgery** | **Internal medicine** |  |
| Total number of patients (capacity) | $N_{pat}$ | 400 | 570 | Data |
| Number of patient profiles | $N_g$ | 1 | | Data |
| Probability of belonging to profile $g$ | $p^g$ | 0.41 | 0.59 | Estimated from data |
| Number of wards | $N_w$ | 10 | 15 | Data |
| Number of distinct procedures | $N_{proc}$ | 15 | | Data |
| Number of device types | $N_k$ | 10 | | Data |
| Profile-specific probability of undergoing procedure $p$ in ward $i$ | $\alpha_{i,p}^g$ | Table S5 | Table S6 | Estimated from data |
| Profile-specific transfer rate between ward $i$ and $j$ | $\tau_{i,j}^g$ | Table S3 | Table S4 | Estimated from data |

**Table 1B.** Pathogen-associated parameters.

| Entry parameters | Notation | Value | Reference |
| --- | --- | --- | --- |
| Profile-specific upon-admission prevalence in ward $w$ | $Prev_w^g$ | Table S1 | Estimated from data |
| Transmission risk for procedure $p$ | $r_p$ | Figure S4 | Henriot *et al.*, 2022 |
| Duration of eclipse phase (min-max) | $(d_{e,min}, d_{e,max})$ | (2-14) (HCV) | Martinello *et al.*, 2018 |
|  |  | (15-15) (HBV) | Candotti et Laperche 2018 |

At each time step, a patient with profile *g* can move from ward *i* to ward *j* with probability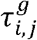. All transfer probabilities are provided in the square transfer matrix 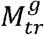 of size (*N*_*w*_*+*1) × (*N*_*w*_*+*1) with the last row being a vector of zeros except at last position (i.e., every discharged patient stays in the “discharged” compartment):

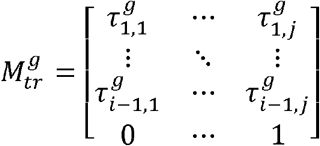

### 2.2 Procedures

In total, *N*_*proc*_ distinct invasive procedures are performed within the hospital. At each time step, a patient of group *g* from ward *i* undergoes procedure *p* with probability 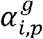. The procedure probability matrix of size *N*_*w*_× (*N*_*proc*_*+*1), is denoted 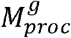. The last column of this matrix is the vector of probabilities associated with the event “no procedure” for each ward (i.e. the patient stays in a given ward without undergoing any procedure):

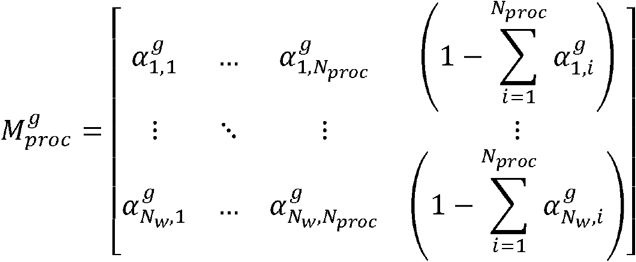

### 2.3 Devices dynamics

Medical or surgical devices are classified into *N*_*k*_ types and associated with procedures by a table giving the device types in the first column and the associated procedures in the other columns. A procedure can be associated with the use of multiple devices.

The quantity of available devices is ward-dependent, type-dependent, and time-dependent. It is divided into 3 device groups: (a) new (sterile) device, (b) previously used uncontaminated device, and (c) previously used contaminated device.

The number of new available devices at time *t* of type *j* in ward *i* is given by 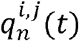; quantities of new devices over the entire hospital are provided in a N_w_ × N_*K*_ sized matrix:

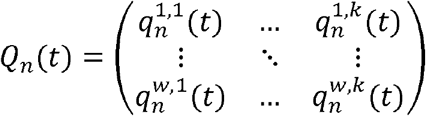

The number of previously used uncontaminated devices at time *t* of type *j* in ward *i* is given by 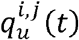 ; quantities of used uncontaminated devices over the entire hospital are provided in a *N*_*w*_× *N*_*K*_ sized matrix:

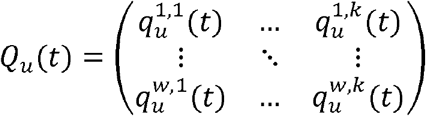

Finally, the number of previously used and contaminated devices at time *t* of type *j* in ward *i* is given by 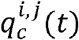 ; quantities of contaminated devices over the entire hospital are provided in a N_w_× N_*K*_ sized matrix:

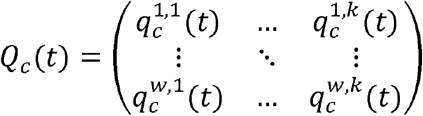

### 2.4 Transmission dynamics

#### 2.4.1 Patients

Upon admission, a patient can be either Susceptible (S) or Infectious (I), assuming that the number of patients entering the hospital shortly after being contaminated (considered as exposed) is negligible. During hospitalization, a Susceptible patient can become Exposed (E) after exposure to a reused contaminated material. We denote *ρ*^*s*^(*t*) the status (i.e., 0 for S, 1 for E, or 2 for I) of patient *s* at time *t*. Initial statuses of patients are randomly drawn according to a vector of upon-admission prevalences giving the initial prevalence (i.e., probability of being pathogen-positive when entering the hospital) for each group of patients.

A device of type *k* is reused on patient *s* at time *t* for procedure *p* in ward *w* if no new device is available in the ward (i.e., 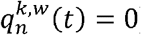). The contamination status of the device is defined by 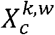, a random variable following a Bernoulli law with 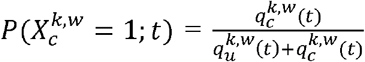 (contaminated device) and 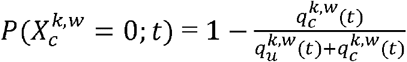 (uncontaminated device).

The time-dependent probability of a susceptible patient getting infected after exposure to reused contaminated devices during procedure *p* in ward *w* is computed as:

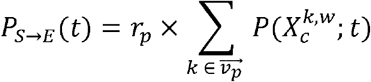

Where 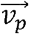 is the vector of device types used during procedure *p* and *r*_*p*_ is the risk of infection after exposure to a contaminated device during the same procedure, randomly drawn in an associated risk distribution.

If infection occurs, the patient becomes exposed at time *t* and his status changes to *ρ*^*s*^(*t*) =1. The patient stays in the exposed state until the end of an eclipse phase *e* is reached. This corresponds to the pre-ramp-up phase of the pathogen natural history during which the patient infectiousness is considered to be null. The eclipse phase duration is drawn in a uniform distribution with parameters *e*_*min*_ and *e*_*max*_ for each new patient *s* entering the hospital:

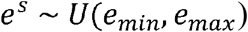

A counter *h*^*s*^(*t*) of the number of time-steps since exposure is initialized upon admission and is given at each time-step by:

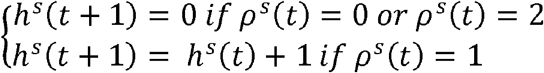

If *h*^*s*^(*t*) = *e*^*s*^, the eclipse phase is over and the exposed patient becomes infectious (^s^) until he leaves the hospital.

#### 2.4.2 Devices

At each time step, in each ward, patients undergo their procedures successively. When 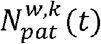 patients in ward *w* need to undergo a procedure requiring a device of type *k*, these procedures requiring the same device type are performed in a random order. The rank of patient *s* is then denoted 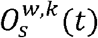 among all procedures requiring device *k* at time *t* in ward *w*.

Any device that is not already contaminated and that is used on an infected individual is considered contaminated after exposure. However, after each procedure, the device undergoes a sterilization process, which successfully clears contamination with probability 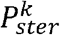 (which is device-specific).

To that aim, we define 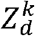 a device-dependent random variable following a Bernoulli law (i.e., device is well disinfected, or insufficiently disinfected) with 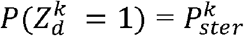 and 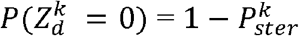.

Between times *t* and *t +1* the dynamics of device type *k* in ward *w* are thus described by the following systems of equation:

If 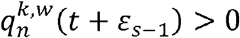 (i.e., new devices of type k are still available in ward w at that time):

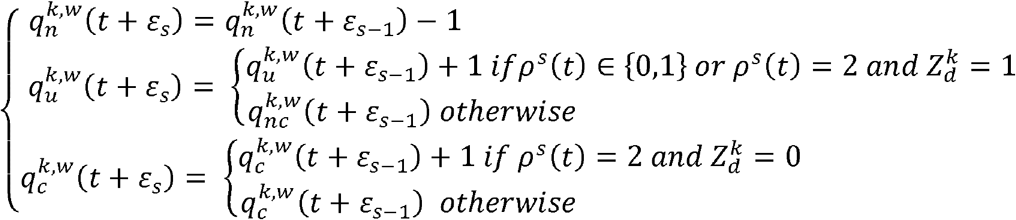

If 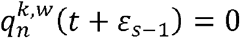 (i.e., no new devices of type k are available in ward w at that time):

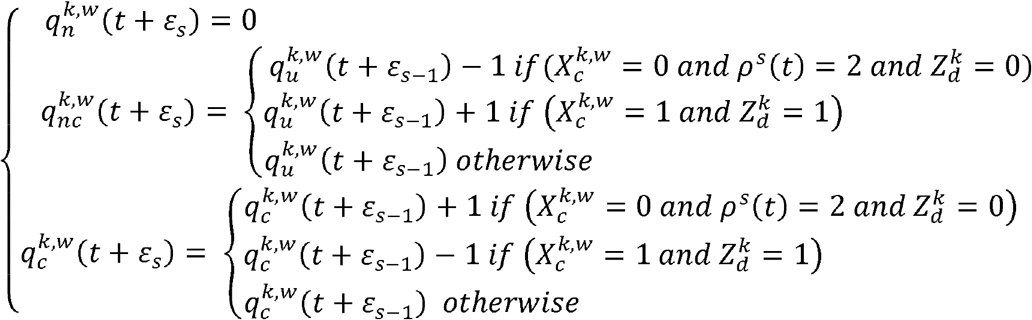

Where 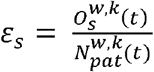 is the increment of time step generated by a procedure using device *k* performed on patient *s* and 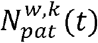 is the total number of patients undergoing a procedure requiring device *k* in ward *w* at time *t*.

Finally, each type of device undergoes a full renewal at a given device-dependent and ward-dependent frequency 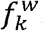. All previously used devices of that type are then replaced by new devices.

### 2.6 Model application

#### 2.6.1 Application to HCV in an Egyptian hospital: baseline scenarios

We used detailed longitudinal data collected in an Egyptian hospital (Ain-Shams University Hospital, Cairo) in 2017 over a 6-month period to inform multiple parameters of our model (ClinicalTrial ID NCT02826447). Five hundred patients were screened upon-admission for HCV positivity and followed over the course of their hospitalization in the Surgery and Internal medicine departments. Many data were collected: (a) Upon-admission patient characteristics such as age, gender and history of previous hospitalization, (b) patient location within the hospital, (c) procedures underwent by these same patients. More details on the data collected is available in Anwar *et al*., 2021 (6).

These data were used to parametrize the model, in particular estimate: (a) The initial prevalence, (b) the number of patients, (c) the number of wards, (d) the number of procedures, (e) the transition matrix between wards within the hospital, and (f) the probability matrix associated with the procedures. These parameters were estimated for both departments (i.e. patient profiles). As the minimum duration of a procedure in our data was 5 minutes, we considered a 5-minute time-step for our transition matrix and procedure probability matrix estimations. In addition, we used other data collected in 2021 to inform the quantity of available devices of each type within this same hospital, the dates of resupply (i.e., renewal frequency), as well as the admission probability in each of the departments. The values of all these parameters and the method used to estimate the resupply dates are available in the supplementary material (Tables S3-S10, Figures S1-S3, Text S1).

Finally, the per-procedure transmission risks were retrieved from a previous work (7) and the minimum and maximum durations of the eclipse phase were found in the literature (8). No information was available to estimate the probability of successful sterilization for each device so these were informed based on expert opinion. As Ain-Shams is a University Hospital, where adherence to control measures is usually high, the probability of successful sterilization was set to 95% for all device types.

In addition to the baseline scenario exploring transmission within a high-resource setting, we explored the case of a hospital with lower resources and lower adherence to control measures. To that aim, we divided the quantity of available supply by two, and the dates of resupply were estimated with this new information. In addition, the probability of successful sterilization was reduced to 80% for all device types.

Our model was run over a year, representing 105,120 time steps. The main outcomes retrieved from our simulations were the daily incidence rate as well as the yearly cumulative incidence at hospital and ward level. In addition, we studied the yearly attributable portion to new HCV cases for each type of device.

#### 2.6.2 Interventions: reinforced infection control

We then simulated two different interventions based on HCV testing to reduce the risk of infection for hospitalized patients:

i. **A targeted ward-level systematic-screening intervention**. The three most at-risk wards (i.e., with the highest yearly estimated cumulative incidence) were identified after running baseline scenarios. Every patient entering one of these three wards was screened for HCV. Reinforced infection control was then implemented for identified positive patients, simulated as systematic successful sterilization of devices following use on these patients (i.e., the probability of successful sterilization was set to 1).
ii. **A random-screening upon admission intervention**. A random subset of patients entering the hospital were screened for HCV. Reinforced infection control was then implemented for identified positive patients, simulated as systematic successful sterilization of devices following use on these patients.

The total number of tests performed in the ward-level intervention over a year was retrieved so that the number of random tests in the random-testing intervention was set to be the same, in order to compare these two scenarios. Test sensitivity and specificity were both assumed to be 100%.

#### 2.6.3 Sensitivity analysis

Various sources of uncertainty might have interfered with the per-procedure risk estimations. In particular, the risk associated with surgery was estimated based on ORs reported in multiple studies (3), but no information was available on whether this procedure was describing the procedure itself or took into account surgeries and other often associated sub-procedures (such as intubation or injection for example) as a whole. Thus, a sensitivity analysis was performed in order to compare our baseline assumption (i.e., that this risk is in fact associated with the act of surgery itself) with the alternative assumption that this risk also accounts for other sub-procedures associated with surgery, in which case we excluded from our database all procedures occurring in the operating room (OR) except the ones reported as surgeries.

#### 2.6.3 Extension to HBV

In order to extend our field of application to the study of HBV transmission in hospitals, we applied our model to the case of an Ethiopian hospital, in which high levels of HBV prevalence were recently found. We assumed a similar hospital structure with three modified input parameters:

i. **The initial prevalence**. We retrieved age-based HBV seroprevalence from Mohammed et al. (2022) (9), who explored HBV infections in two Ethiopian hospitals.
ii. **The per-procedure infection risks**. We estimated per-procedure infection risk distributions for HBV by multiplying those computed for HCV by the ratio of the probability of HBV infection after exposure to contaminated blood (6-30%) over the same probability for HCV (1.8%) (10).
iii. **The duration of the eclipse phase**. From the literature, we estimated the eclipse phase duration for HBV to be 15 days (11)

More information on these calculations is available in Supplementary Material. All other parameters were kept unchanged. We explored the same scenarios than for HCV: high-resource and low-resource hospitals.

#### 2.6.6 Model simulations

This model was coded in C++ using the *Rcpp* interface in R version 4.3.1 (12). All simulations were performed using the same R version. The number of simulations needed to catch most of the variability produced by the model was estimated by studying the convergence of the cumulative mean of the number of yearly number of cases for the low-resource setting scenario. Figure S8 shows that the mean converges after approximately 100 simulations. Therefore, each scenario was simulated 100 times.

## 3 Results

### 3.1 HCV in Egypt

#### 3.1.1 Baseline scenarios

The predicted yearly number of HCV acquisitions among hospitalized patients differed highly depending on the scenario.

In the high-resource setting case, this number was estimated around 4.6 cases per 100,000 patient/year (95% PI [0.6-9.5]) (Fig 2A). The yearly incidence rate was the highest in the OR (19.3 cases per 100,000 per year, 95% PI [0-40.4]), emergency room intensive-care unit (ER ICU) (4.5 cases per 100,000 per year, 95% PI [0-88.8]) and Ophthalmology (1.1 cases per 100,000 per year, 95% PI [0-8.3]) wards (Fig 2B), though leading to less than 3, 0.5 and 0.14 annual cases on average, respectively. The yearly cumulative incidence was higher in the surgery department (7.4 cases per 100,000 per year, 95% PI [0-15.2]) compared to the internal medicine department (0.4 cases per 100,000 per year, 95% PI [0-1.8]). The type of device associated with the largest number of HCV contaminations was the endotracheal tube, with 90% of all cases on average (Fig 3A).

**Figure 2.**
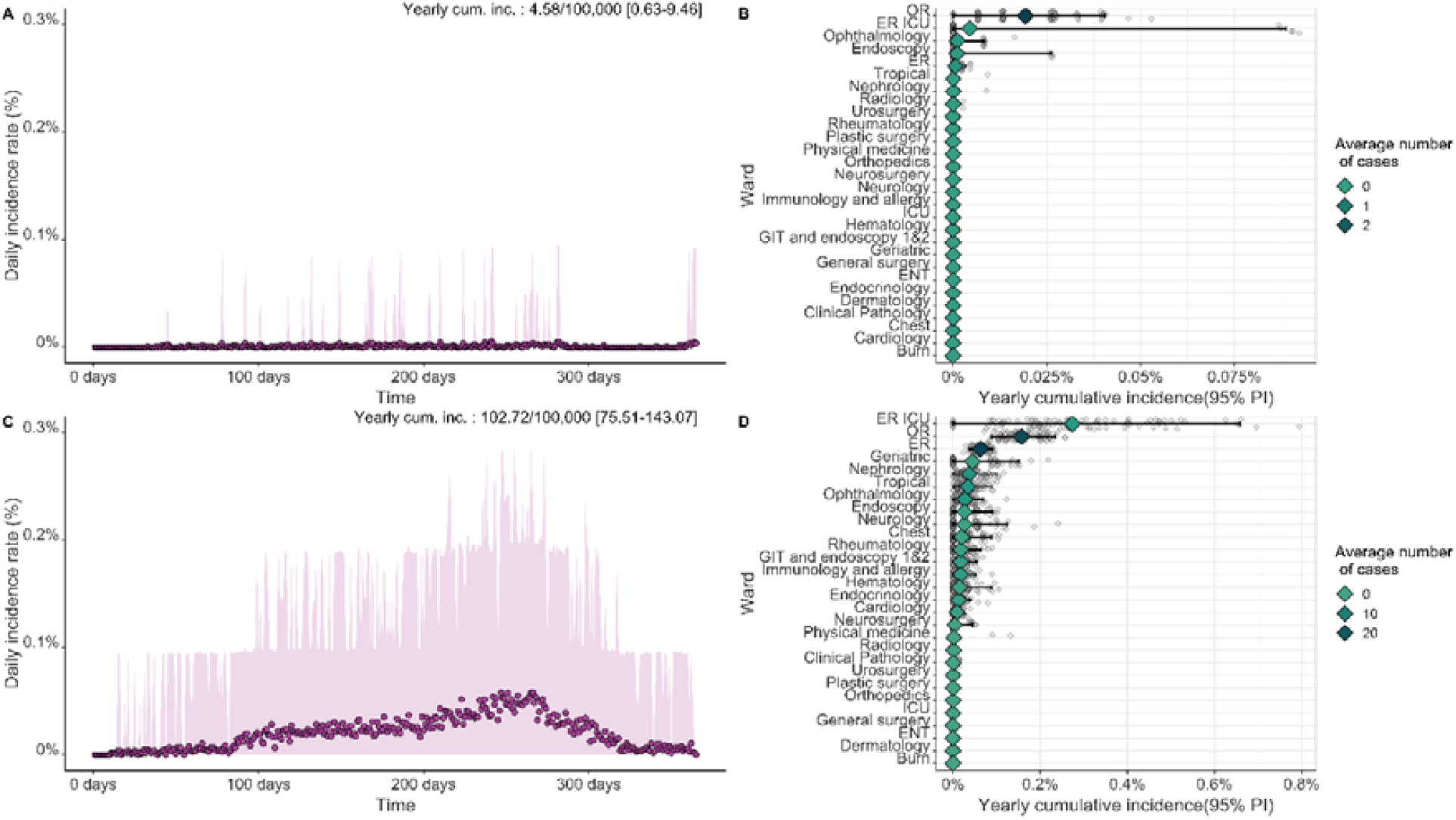
Results of the model for baseline scenarios (HCV case). (A) and (C): Daily incidence rate for the high-resource hospital and low-resource hospital, respectively. (B) and (D) Yearly cumulative incidence (mean and 95% PI) and average number of cases for each ward, ranked by mean cumulative incidence values.

**Figure 3.**
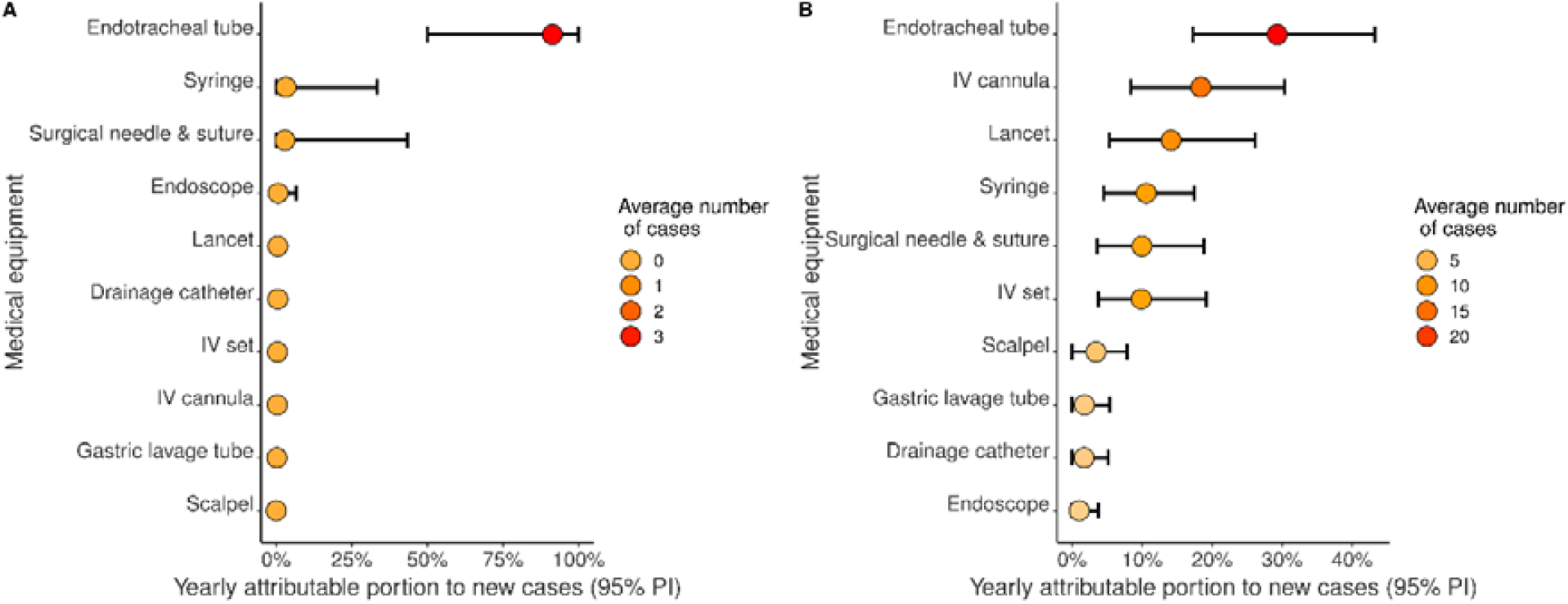
Yearly portion of new HCV cases attributable to each device, in (A) the baseline scenario for the high-resource hospital and (B) the baseline scenario for the lowresource hospital.

In the low-resource setting case, the average yearly number of cases was, as expected, much higher, estimated at 102.7 per 100,000 patient/year (95% PI [75.5-143.1]) (Fig 2C). The ER ICU, OR and ER wards were associated with the highest yearly incidence, with 273.6 cases per 100,000 patient/year (95% PI [0-658.9]), 157.8 cases per 100,000 patient/year (95% PI [88.6-235.3]) respectively (Fig 2D). Again, the yearly cumulative incidence was higher in the surgery department (60.8 cases per 100,000 per year, 95% PI [38.1-88.8]) compared to the internal medicine department (43.3 cases per 100,000 per year, 95% PI [22.9-74.4]). Here also, endotracheal tubes were associated with the highest number of HCV contaminations, representing almost 30% of all HCV infections on average, followed by intravenous (IV) canulas (around 15% of all infection cases) (Fig 3B).

#### 3.1.2 Interventions

In each case (i.e. high-resource or low-resource setting), the efficacy of the targeted wardlevel systematic screening interventions was found to be superior to that of the uponadmission random-testing interventions (Figure 4).

**Figure 4.**
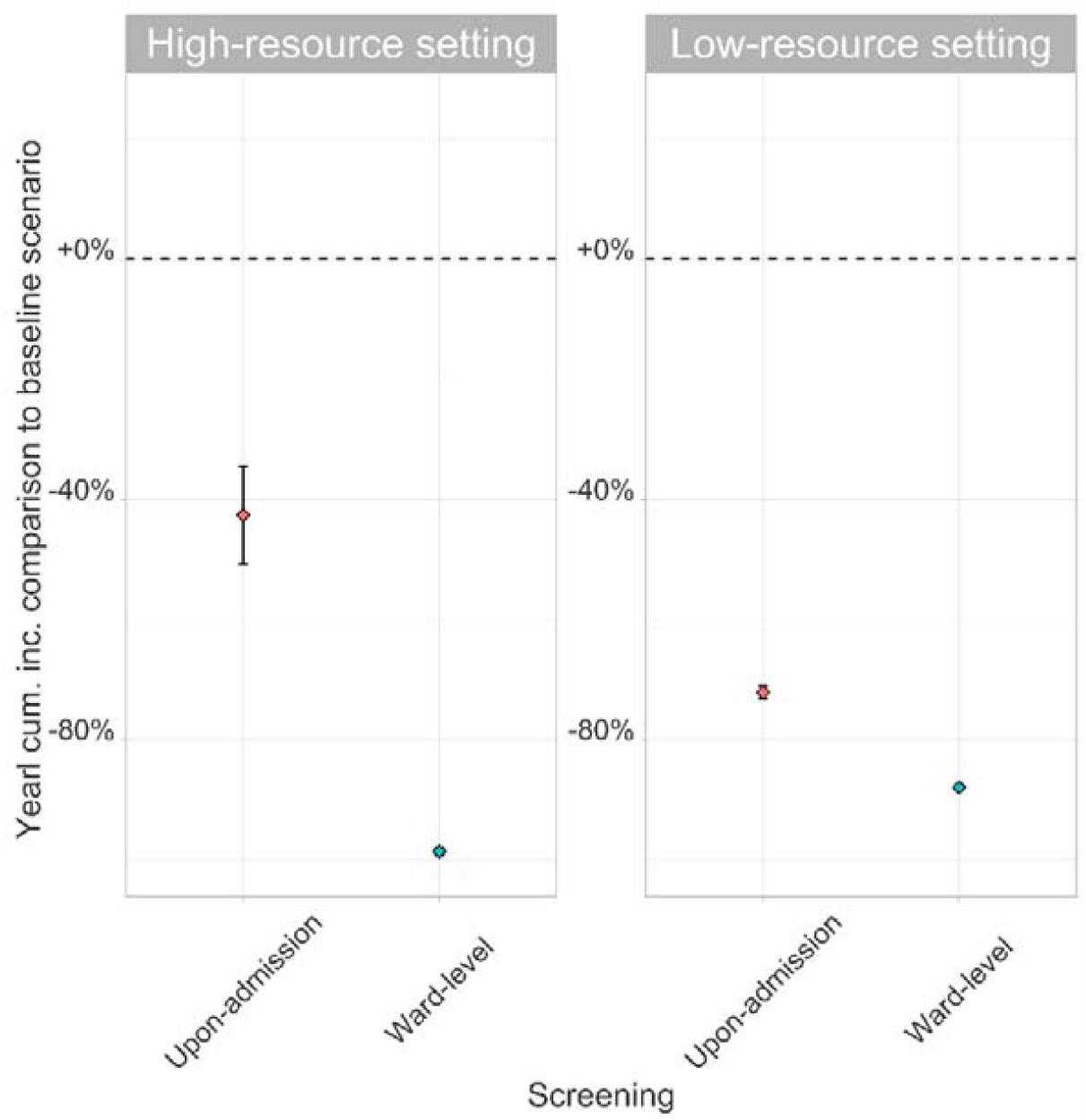
Yearly cumulative incidence comparison to baseline scenarios for two different intervention strategies (95% CI of the mean) for (A) high-resource and (B) low-resource hospitals. Baseline scenarios corresponds to the no-intervention scenarios. For the highresource and low-resource settings on average of 30,375 patients (40.5%) and 53,472 (71%) patients were screened, respectively, (i) either systematically (in the three most at-risk wards) or (ii) randomly upon admission.

For the high-resource setting, a total average number of 30,375 patients (40.5%) went through the OR, ER ICU or Ophthalmology wards during their hospitalization. Performing systematic screening in these three most at-risk wards reduced the annual number of HCV acquisition cases by 97.6% (95% CI [96.5-98.4]) on average compared to the baseline scenario, whereas random testing in 40.5% of patients upon admission reduced the annual number of cases by an average of 41.6% (95% CI [37.6-45.5]).

Trends were similar for the low-resource setting case. The three most at-risk wards were the ER ICU, OR, and ER wards, representing an average of 53,472 patients (71%). Performing systematic screening in these wards allowed a reduction of 88.5% (95% CI [88.0-89.0]). Performing random screening in 71% of newly admitted patients reduced the annual number of cases by an average of 72.1% (95% CI [71.4-72.7]).

#### 3.1.3. Sensitivity analysis

When considering that our estimate of HCV infection risk associated with surgery already takes into account the risk of other associated sub-procedures, the yearly cumulative incidence was reduced by 73.7% in the high-resource setting baseline scenario (with 1.2 cases per 100,000 patient/year ; 95% PI [0-3.7]) and by 22.9% in the low-resource setting baseline scenario (79.2 cases per 100,000 patient/year ; 95% PI [52.2-117.9]), compared to the baseline case where each of the procedures occurring in the operating room was taken into account individually.

### 3.2 HBV in Ethiopia

#### 3.2.1 Baseline scenarios

The predicted number of HBV acquisitions among hospitalized patients was higher than for HCV, and again differed highly between low- and high-infection control hospital settings (Fig. S5).

In the high-resource setting, this number was estimated at 49.8 cases per 100,000 patient/year (95% PI [38.2-70.2]), with the OR ward being the most at-risk, with a yearly cumulative incidence of 210 cases per 100,000 patient/year (95% PI [160-291.2]), followed by the ER ICU ward (17 cases per 100,000 patient/year, 95% PI [0-91]) and the Endoscopy ward (11.5 cases per 100,000 patient/year, 95% PI [0-47.9]). The type of device associated with highest number of HCV contaminations was the endotracheal tube with more than 80% of all cases on average (Fig S6).

In the low-resource setting, the yearly cumulative incidence reached 860.8 cases per 100,000 patient/year (95% PI [667.8-1119]), with the ER ICU ward again being one of the most at-risk with a yearly cumulative incidence of 1823.9 cases per 100,000 patient/year (95% PI [533-3635]), followed by the OR ward (1608.3 cases per 100,000 patient/year, 95% PI [1122.5-2288.6]), and the ER ward (492.6 cases per 100,000 patient/year, 95% PI [388.2-628.1]). Again, the endotracheal tube was associated with the highest number of contaminations, accounting for more than 30% of all contaminations, on average (Fig S6).

## 4. Discussion

Herein we describe the framework of a novel agent-based model of the nosocomial transmission of bloodborne pathogens. Using data on patient movements within the hospital, procedures underwent by patients and the hospital structure, we were able to reproduce the dynamics of patients and medical devices within the hospital. We showed that the risk of HCV infection is low in a hospital with high resources and good disinfection practices (less than 5 cases per 100,000 patients per year) but higher for a low-resource hospital with around 100 cases per 100,000 patients per year. For the HBV case, the trends were the same but with higher absolute risk values.

Until now, and to our knowledge, no such model was available in the literature and all previous modelling approaches to explore bloodborne pathogen spread in healthcare settings were based on quantitative risk assessment (13,14). Dynamic models had already been used to study the transmission dynamics of HCV and other bloodborne pathogens in the community, especially within IDU networks as reviewed by Cousien et al. (2015) (5). However, most of these models were fully compartmental and only few used an individual-based approach.

The community incidence of HCV in rural villages in Egypt was estimated at 37/100,000 per year in 2018, soon after the implementation of a mass HCV screening campaign. Even if this may not be fully comparable with our results, our estimations remain in the same order of magnitude (15). Our work suggests that hospitals might still generate new HCV and HBV contaminations, especially in healthcare settings with low resources and/or suboptimal disinfection practices. This is in line with recent studies highlighting a persisting increased risk of HBV and HCV infection associated with medical and surgical procedures (2,16). We showed that nosocomial acquisitions could however be tackled by improving control measures and by a better allocation of financial resources to make more sterile devices available for hospitals.

Some limitations related to the model structure and data used to inform the model may be highlighted.

First, the model only accounts for between-patient transmission without considering transmission from healthcare workers to patients or from patients to healthcare workers. Cases of contamination through these routes have been reported, leading us to potentially underestimate acquisition risks. However, such cases are usually quite rare (17).

Second, our results highly depend on the assumed per-procedure risks of transmission through blood contact. However, as mentioned in the method section, these risks might have been over-or under-estimated. Our sensitivity analysis showed that the estimated yearly cumulative incidence could vary depending on the assumption made on the risk associated with surgery: considering that this risk takes into account the risk of other associated sub-procedures led to a lower yearly cumulative incidence.

Third, our model allowed us to assess the effectiveness of ward-level targeted systematic and upon-admission random testing in the hospital. While randomly testing patients upon-admission appeared to be a sub-optimal strategy, our results suggested that systematically screening patients admitted into the three most at-risk wards could be more efficient in reducing the yearly number of cases. Nevertheless, we assumed perfect disinfection of devices used on positive patients, which might be difficult to reach in practice. Other interventions could be assessed using our model. In particular, a more realistic intervention would be device reallocation between wards, which could reduce the overall number of infections by setting the risk to zero in most at-risk wards while potentially increasing it in other wards.

Fourth, the data used to inform the quantity of available devices within wards was approximated using the available devices at the entire Ain Shams hospital level, assuming that these devices were allocated to wards proportionally to admissions. This may not reflect the real supply allocation. In addition, we assumed that the available quantity of devices in low-resource hospitals was twice lower than for high-resource hospitals, without any data to base this on. Finally, probabilities of successful disinfection were chosen based on expert opinion but do not rely on data collected within hospitals.

Many of these limitations stem from a lack of data to correctly inform the corresponding model parameters, forcing us to make assumptions. However, our aim here was mostly to describe a new modelling framework and to illustrate its potential applications, rather than to directly inform public health decision-making regarding HCV or HBV control in hospitals.

On a related note, our results may not accurately describe the current Egyptian situation as we used data from 2017. The current upon-admission prevalence of HCV infected patients might be much lower, as a massive test and treat program launched by the Egyptian government in 2018 led to the treatment of more than 2 million Egyptian and the national prevalence in 2023 is expected to fall around 0.5% (18).

Nevertheless, some countries still report a high HCV or HBV prevalence. A recent meta-analysis focusing on HBV prevalence in sub-continental countries showed that Pakistan and India still have a national prevalence over 5% (19). In addition, nosocomial outbreaks of bloodborne infections still occur nowadays, even in developed countries. Between 2006 and 2020, 91 outbreaks of transmission of HBV and HCV within hospitals have been reported in Europe, corresponding to 442 cases of infections (20). In the USA, such events were reported 66 times between 2008 and 2019, corresponding to more than 500 new infections (21). Our model could help predict such outbreaks, track past contamination events and assess the effectiveness of intervention measures.

To conclude, the modelling tool we propose may be useful to study the spread of bloodborne pathogens at a hospital level and assess the efficacy of multiple interventions on the reduction of their transmission, as well as the associated costs. It could help implement more efficient prevention measures in hospitals and, for instance, make WHO HCV elimination targets more easily achieved. The advantage of this model lies in its high flexibility, while computation times remain reasonable despite model complexity. It can account for heterogeneity between patient profiles by informing different transition matrices and could be easily applied to other hospitals in other contexts beyond those described here.It model could also help study the transmission of other existing bloodborne pathogens such as Zika virus, or prions, which are yet poorly understood. In addition, as experienced with the COVID-19 pandemic, emerging pathogens can lead to a huge public health burden. Being able to model the transmission of potential emerging bloodborne pathogens could help predict the associated disease burden in hospitals and help implement efficient public health policies.

## Data Availability

The data analysed in this study is available upon request only. Individual data requests may be sent to the CorC.

