## Supplementary_Appendix for "An agent-based model to simulate the transmission dynamics of bloodborne pathogens within hospitals"

**Table S1.** Ward ID and names………………………………………………………………………………………….2

**Table S2.** Matrix of association between devices and procedures……………………………………………………………………………3

**Text S1.** Method used to estimate the resupply frequency…………………………………………………………………………………….…3

**Table S3.** Transition matrix for patients hospitalized in the surgery department…………………………………………………..…4

**Table S4.** Transition matrix for patients hospitalized in the internal medicine department…………………………..…………5

**Table S10.** Yearly initial quantity of previously used devices in each ward for the low-resource setting ……………..……9

**Figure S1.** Use of devices over available quantity for the high-resource setting……………………………………………….....…10

**Figure S2.** Use of devices over available quantity for the low-resource setting………………………………………...…………….10

**Figure S3.** Yearly equipment uses in each ward. …………………………………………………………………………………………….………11

**Figure S4.** Distribution of the risk of HCV infection associated with different procedures…………………………………….…11

**Figure S5.** Results of the model for baseline scenarios (HBV case). …………………………………………………………….…………12

**Figure S6.** Yearly attributable portion to new cases for each device (HBV case)…………………….………………………………12

**Figure S7.** Yearly cumulative incidence for two different intervention strategies (HBV case) ……………………..…………13

**Figure S8.** Cumulative mean of the number of annual cases for 1 to 200 simulations. ……………………………..……………13

**Table S1.** Ward ID and names

| **Ward ID** | **Ward name** | **Initial prevalence (HCV)** | **Initial prevalence (HBV)** |
| --- | --- | --- | --- |
| 1 | GIT & Endoscopy | 0% | 0% |
| 2 | Endocrinology | 50% | 11.5% |
| 3 | Immunology & Allergy | 0% | 0% |
| 4 | Hematology | 0% | 7.1% |
| 5 | Rheumatology | 0% | 9.3% |
| 6 | Geriatric | 0% | 0% |
| 7 | Chest | 0% | 6% |
| 8 | Nephrology | 0% | 3% |
| 9 | Cardiology | 0% | 13% |
| 11 | Neurology | 0% | 14% |
| 12 | Tropical medicine | 51.8% | 8.4% |
| 13 | Dermatology | 0% | 0% |
| 14 | Ophtalmology | 13.6% | 7.5% |
| 16 | ICU | 0% | 0% |
| 17 | Orthopedics | 3.1% | 7.4% |
| 18 | General surgery | 4.2% | 7.4% |
| 20 | Neurosurgery | 25% | 9% |
| 21 | Urosurgery | 100% | 6.3% |
| 22 | Burn | 0% | 0% |
| 23 | ENT | 0% | 0% |
| 24 | Plastic surgery | 5.3% | 6.2% |
| 25 | Operating room (OR) | 0% | 0% |
| 26 | Radiology | 0% | 0% |
| 27 | Clinical pathology | 0% | 3% |
| 28 | Endoscopy | 0% | 0% |
| 29 | Physical medicine | 0% | 9.5% |
| 30 | Emergency room (ER) | 9% | 7.3% |
| 31 | Emergency room ICU | 0% | 0% |

**Table S2.** Matrix of association between devices and procedures. Each one represents an association between a procedure and a device.

| Procedure  Device | Surgery | IV catheter | Suture | Blood transfusion | Blood sample | Injection | Endoscopy | Gastric lavage | Cardiac catheter | Dialysis | Wound dressing | Blood glucose | Endotracheal intubation |
| --- | --- | --- | --- | --- | --- | --- | --- | --- | --- | --- | --- | --- | --- |
| Syringes | 1 | 0 | 0 | 0 | 1 | 1 | 0 | 0 | 0 | 0 | 0 | 0 | 0 |
| IV Set | 1 | 1 | 0 | 1 | 0 | 0 | 0 | 0 | 1 | 1 | 0 | 0 | 0 |
| IV Cannula | 1 | 1 | 0 | 1 | 0 | 0 | 0 | 0 | 1 | 1 | 0 | 0 | 0 |
| Scalpel | 1 | 0 | 0 | 0 | 0 | 0 | 0 | 0 | 0 | 0 | 0 | 0 | 0 |
| Lancet | 0 | 0 | 0 | 0 | 0 | 0 | 0 | 0 | 0 | 0 | 0 | 1 | 0 |
| Surgical needles & suture kits | 1 | 0 | 1 | 0 | 1 | 0 | 0 | 0 | 0 | 0 | 1 | 0 | 0 |
| Endotracheal tube | 1 | 0 | 0 | 0 | 0 | 0 | 0 | 0 | 0 | 0 | 0 | 0 | 1 |
| Drainage catheter | 0 | 0 | 0 | 0 | 0 | 0 | 0 | 0 | 0 | 0 | 0 | 0 | 0 |
| Gastric lavage tube | 0 | 0 | 0 | 0 | 0 | 0 | 0 | 1 | 0 | 0 | 0 | 0 | 0 |
| Endoscope | 0 | 0 | 0 | 0 | 0 | 0 | 1 | 0 | 0 | 0 | 0 | 0 | 0 |

**Text S1.** Method used to estimate the resupply frequency

The date of device renewal ($t_{\mu}$) was calculated based on the available quantity of a given device for one year ($Q_{tot}$) and the per-time-step quantity use of this same device (n). It was defined so that the initial quantity of new device for year 1 is equal to the initial quantity of new device for year 2.

We define r, the ratio of the available device quantity over the yearly usage: $r=Q_{tot}/(n\times t_{max}$)

We can infer the following device use :

- From $t_{0}$ to $t_{\emptyset}$ : $Q_{tot}\times r$
- From $t_{\mu}$ to $t_{max}$ : $Q_{tot}\times(1-r$)
- From $t_{\emptyset}$ to $t_{\mu}$ : $(n\times t_{max}$)$-Q_{tot}$

Finally, we have $\boldsymbol{t}_{\boldsymbol{\mu}}\boldsymbol{=}\frac{\boldsymbol{Q}_{\boldsymbol{tot}}\boldsymbol{\times}\left( \boldsymbol{r-1} \right)\boldsymbol{+(n\times}\boldsymbol{t}_{\boldsymbol{max}}\mathbf{)}}{\boldsymbol{n}}$

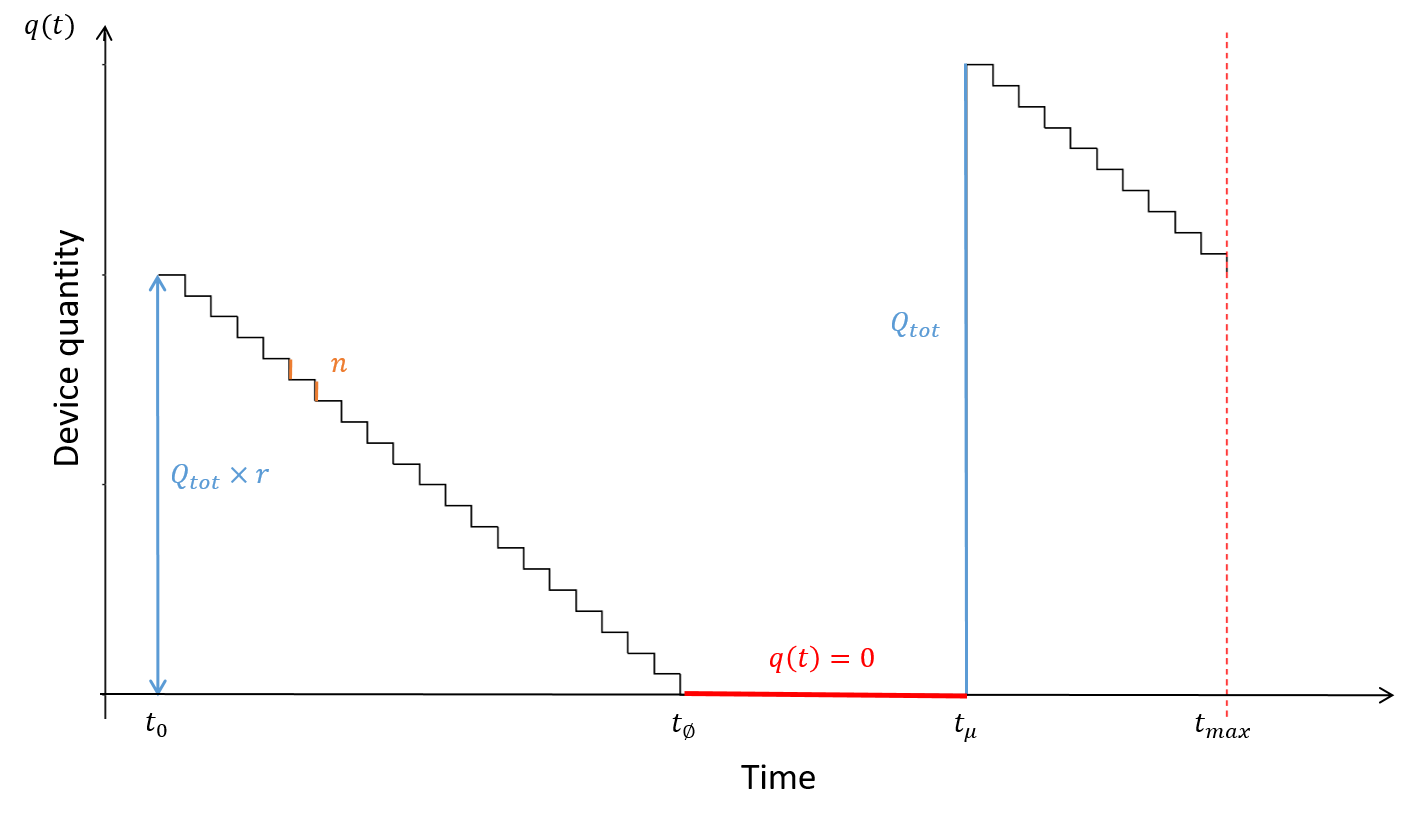

**Table S3.** Transition matrix for patients hospitalized in the surgery department

|  | **1** | **2** | **3** | **4** | **5** | **6** | **7** | **8** | **9** | **11** | **12** | **13** | **14** | **16** | **17** | **18** | **20** | **21** | **22** | **23** | **24** | **25** | **26** | **27** | **28** | **29** | **30** | **31** | **out** |
| --- | --- | --- | --- | --- | --- | --- | --- | --- | --- | --- | --- | --- | --- | --- | --- | --- | --- | --- | --- | --- | --- | --- | --- | --- | --- | --- | --- | --- | --- |
| **1** | 0 | 0 | 0 | 0 | 0 | 0 | 0 | 0 | 0 | 0 | 0 | 0 | 0 | 0 | 0 | 0 | 0 | 0 | 0 | 0 | 0 | 0 | 0 | 0 | 0 | 0 | 0 | 0 | 0 |
| **2** | 0 | 0 | 0 | 0 | 0 | 0 | 0 | 0 | 0 | 0 | 0 | 0 | 0 | 0 | 0 | 0 | 0 | 0 | 0 | 0 | 0 | 0 | 0 | 0 | 0 | 0 | 0 | 0 | 0 |
| **3** | 0 | 0 | 0 | 0 | 0 | 0 | 0 | 0 | 0 | 0 | 0 | 0 | 0 | 0 | 0 | 0 | 0 | 0 | 0 | 0 | 0 | 0 | 0 | 0 | 0 | 0 | 0 | 0 | 0 |
| **4** | 0 | 0 | 0 | 0 | 0 | 0 | 0 | 0 | 0 | 0 | 0 | 0 | 0 | 0 | 0 | 0 | 0 | 0 | 0 | 0 | 0 | 0 | 0 | 0 | 0 | 0 | 0 | 0 | 0 |
| **5** | 0 | 0 | 0 | 0 | 0 | 0 | 0 | 0 | 0 | 0 | 0 | 0 | 0 | 0 | 0 | 0 | 0 | 0 | 0 | 0 | 0 | 0 | 0 | 0 | 0 | 0 | 0 | 0 | 0 |
| **6** | 0 | 0 | 0 | 0 | 0 | 0 | 0 | 0 | 0 | 0 | 0 | 0 | 0 | 0 | 0 | 0 | 0 | 0 | 0 | 0 | 0 | 0 | 0 | 0 | 0 | 0 | 0 | 0 | 0 |
| **7** | 0 | 0 | 0 | 0 | 0 | 0 | 0 | 0 | 0 | 0 | 0 | 0 | 0 | 0 | 0 | 0 | 0 | 0 | 0 | 0 | 0 | 0 | 0 | 0 | 0 | 0 | 0 | 0 | 0 |
| **8** | 0 | 0 | 0 | 0 | 0 | 0 | 0 | 0 | 0 | 0 | 0 | 0 | 0 | 0 | 0 | 0 | 0 | 0 | 0 | 0 | 0 | 0 | 0 | 0 | 0 | 0 | 0 | 0 | 0 |
| **9** | 0 | 0 | 0 | 0 | 0 | 0 | 0 | 0 | 0,875 | 0 | 0 | 0 | 0 | 0 | 0,09375 | 0,03125 | 0 | 0 | 0 | 0 | 0 | 0 | 0 | 0 | 0 | 0 | 0 | 0 | 0 |
| **11** | 0 | 0 | 0 | 0 | 0 | 0 | 0 | 0 | 0 | 0 | 0 | 0 | 0 | 0 | 0 | 0 | 0 | 0 | 0 | 0 | 0 | 0 | 0 | 0 | 0 | 0 | 0 | 0 | 0 |
| **12** | 0 | 0 | 0 | 0 | 0 | 0 | 0 | 0 | 0 | 0 | 0 | 0 | 0 | 0 | 0 | 0 | 0 | 0 | 0 | 0 | 0 | 0 | 0 | 0 | 0 | 0 | 0 | 0 | 0 |
| **13** | 0 | 0 | 0 | 0 | 0 | 0 | 0 | 0 | 0 | 0 | 0 | 0 | 0 | 0 | 0 | 0 | 0 | 0 | 0 | 0 | 0 | 0 | 0 | 0 | 0 | 0 | 0 | 0 | 0 |
| **14** | 0 | 0 | 0 | 0 | 0 | 0 | 0 | 0 | 0 | 0 | 0 | 0 | 0 | 0 | 0 | 0 | 0 | 0 | 0 | 0 | 0 | 0 | 0 | 0 | 0 | 0 | 0 | 0 | 0 |
| **16** | 0 | 0 | 0 | 0 | 0 | 0 | 0 | 0 | 0 | 0 | 0 | 0 | 0 | 0,998093 | 0,000636 | 0,000318 | 0,000636 | 0 | 0 | 0 | 0 | 0 | 0,000318 | 0 | 0 | 0 | 0 | 0 | 0 |
| **17** | 0 | 0 | 0 | 0 | 0 | 0 | 0 | 0 | 3,50E-05 | 0 | 0 | 0 | 0 | 0 | 0,997379 | 0,000104 | 2,30E-05 | 0 | 0 | 0 | 2,30E-05 | 0,000543 | 0,001028 | 0,000115 | 0 | 0 | 5,80E-05 | 0 | 0,000693 |
| **18** | 0 | 0 | 0 | 0 | 0 | 0 | 0 | 0 | 9,00E-06 | 0 | 0 | 0 | 0 | 0 | 4,30E-05 | 0,996972 | 1,70E-05 | 9,00E-06 | 0 | 0 | 1,70E-05 | 0,000981 | 0,00037 | 0,000146 | 1,70E-05 | 0 | 0 | 0 | 0,001419 |
| **20** | 0 | 0 | 0 | 0 | 0 | 0 | 0 | 0 | 0 | 0 | 0 | 0 | 0 | 0,000107 | 0,000107 | 0 | 0,997964 | 0 | 0 | 0,000107 | 0 | 0,000429 | 0,000536 | 0,000107 | 0 | 0 | 0 | 0 | 0,000643 |
| **21** | 0 | 0 | 0 | 0 | 0 | 0 | 0 | 0 | 0 | 0 | 0 | 0 | 0 | 0 | 0 | 0 | 0 | 0,997276 | 0 | 0 | 0 | 0,000227 | 0,000908 | 0 | 0 | 0 | 0 | 0 | 0,001589 |
| **22** | 0 | 0 | 0 | 0 | 0 | 0 | 0 | 0 | 0 | 0 | 0 | 0 | 0 | 0 | 0,035714 | 0 | 0 | 0 | 0,928571 | 0 | 0 | 0 | 0,035714 | 0 | 0 | 0 | 0 | 0 | 0 |
| **23** | 0 | 0 | 0 | 0 | 0 | 0 | 0 | 0 | 0 | 0 | 0 | 0 | 0 | 0 | 0 | 0,047619 | 0 | 0 | 0 | 0,952381 | 0 | 0 | 0 | 0 | 0 | 0 | 0 | 0 | 0 |
| **24** | 0 | 0 | 0 | 0 | 0 | 0 | 0 | 0 | 0 | 0 | 0 | 0 | 0 | 0 | 4,60E-05 | 4,60E-05 | 0 | 0 | 0 | 0 | 0,996007 | 0,001393 | 0,000325 | 0,000557 | 0 | 0 | 0 | 0 | 0,001625 |
| **25** | 0 | 0 | 0 | 0 | 0 | 0 | 0 | 0 | 0 | 0 | 0 | 0 | 0 | 0,000421 | 0,006453 | 0,016552 | 0,000421 | 0,00014 | 0 | 0 | 0,004208 | 0,971665 | 0 | 0 | 0 | 0 | 0 | 0 | 0,00014 |
| **26** | 0 | 0 | 0 | 0 | 0 | 0 | 0 | 0 | 0 | 0 | 0 | 0 | 0 | 0,000759 | 0,040971 | 0,021624 | 0,001897 | 0,001517 | 0,000379 | 0 | 0,00569 | 0,001138 | 0,901745 | 0,000759 | 0 | 0 | 0,02352 | 0 | 0 |
| **27** | 0 | 0 | 0 | 0 | 0 | 0 | 0 | 0 | 0 | 0 | 0 | 0 | 0 | 0 | 0,018519 | 0,034722 | 0 | 0 | 0,002315 | 0 | 0,020833 | 0 | 0,030093 | 0,837963 | 0 | 0 | 0,055556 | 0 | 0 |
| **28** | 0 | 0 | 0 | 0 | 0 | 0 | 0 | 0 | 0 | 0 | 0 | 0 | 0 | 0 | 0 | 0,022989 | 0 | 0 | 0 | 0 | 0 | 0 | 0 | 0 | 0,977011 | 0 | 0 | 0 | 0 |
| **29** | 0 | 0 | 0 | 0 | 0 | 0 | 0 | 0 | 0 | 0 | 0 | 0 | 0 | 0 | 0 | 0 | 0 | 0 | 0 | 0 | 0 | 0 | 0 | 0 | 0 | 0 | 0 | 0 | 0 |
| **30** | 0 | 0 | 0 | 0 | 0 | 0 | 0 | 0 | 0 | 0 | 0 | 0 | 0 | 0 | 0,004843 | 0,017528 | 0,000231 | 0,000461 | 0 | 0 | 0,002076 | 0,000692 | 0,02214 | 0,006227 | 0 | 0 | 0,945803 | 0 | 0 |
| **31** | 0 | 0 | 0 | 0 | 0 | 0 | 0 | 0 | 0 | 0 | 0 | 0 | 0 | 0 | 0 | 0 | 0 | 0 | 0 | 0 | 0 | 0 | 0 | 0 | 0 | 0 | 0 | 0 | 0 |
| **out** | 0 | 0 | 0 | 0 | 0 | 0 | 0 | 0 | 0 | 0 | 0 | 0 | 0 | 0 | 0 | 0 | 0 | 0 | 0 | 0 | 0 | 0 | 0 | 0 | 0 | 0 | 0 | 0 | 1 |

|  | **1** | **2** | **3** | **4** | **5** | **6** | **7** | **8** | **9** | **11** | **12** | **13** | **14** | **16** | **17** | **18** | **20** | **21** | **22** | **23** | **24** | **25** | **26** | **27** | **28** | **29** | **30** | **31** | **out** |
| --- | --- | --- | --- | --- | --- | --- | --- | --- | --- | --- | --- | --- | --- | --- | --- | --- | --- | --- | --- | --- | --- | --- | --- | --- | --- | --- | --- | --- | --- |
| **1** | 0,997737 | 0,000112 | 2,80E-05 | 0 | 0 | 0,000112 | 0 | 0,000251 | 0,000168 | 0 | 0 | 0 | 5,60E-05 | 0 | 0 | 0 | 0 | 0 | 0 | 0 | 0 | 0 | 0,000698 | 0 | 0,00014 | 0 | 0 | 2,80E-05 | 0,000671 |
| **2** | 6,20E-05 | 0,997485 | 6,20E-05 | 0 | 4,10E-05 | 0 | 2,10E-05 | 0,000144 | 0,000247 | 8,20E-05 | 4,10E-05 | 0 | 2,10E-05 | 0 | 0 | 2,10E-05 | 0 | 0 | 0 | 0 | 0 | 0 | 0,000928 | 0 | 0,000124 | 0 | 2,10E-05 | 0 | 0,000701 |
| **3** | 2,20E-05 | 8,80E-05 | 0,997426 | 2,20E-05 | 4,40E-05 | 4,40E-05 | 0 | 0,000242 | 0,000396 | 0 | 0 | 4,40E-05 | 6,60E-05 | 0 | 0 | 4,40E-05 | 0 | 0 | 0 | 0 | 0 | 0 | 0,000902 | 0 | 6,60E-05 | 0 | 6,60E-05 | 2,20E-05 | 0,000506 |
| **4** | 0 | 0 | 5,40E-05 | 0,998757 | 0 | 0 | 0 | 0 | 0 | 0 | 0 | 0 | 0 | 0 | 0 | 0 | 0 | 0 | 0 | 0 | 0 | 0 | 0,000756 | 0 | 0 | 0 | 0 | 5,40E-05 | 0,000378 |
| **5** | 0 | 5,10E-05 | 5,10E-05 | 0 | 0,997434 | 0 | 0 | 0,000154 | 0,000462 | 5,10E-05 | 0 | 0 | 0,000205 | 0 | 0 | 0 | 0 | 0 | 0 | 0 | 0 | 5,10E-05 | 0,000873 | 0 | 0 | 0 | 0 | 0 | 0,000667 |
| **6** | 0,000338 | 8,40E-05 | 0 | 0 | 0 | 0,998649 | 0 | 0,000169 | 0,000169 | 0 | 0 | 0 | 0 | 0 | 0 | 0 | 0 | 0 | 0 | 0 | 0 | 0 | 0,000253 | 0 | 8,40E-05 | 0 | 0 | 0 | 0,000253 |
| **7** | 0 | 0 | 0 | 0 | 0 | 0 | 0,998868 | 0 | 0,000113 | 0 | 0 | 0 | 0,000113 | 0 | 0 | 0 | 0 | 0 | 0 | 0 | 0 | 0 | 0,000566 | 0 | 0 | 0 | 0 | 0 | 0,00034 |
| **8** | 0,000184 | 0,00021 | 0,000315 | 2,60E-05 | 7,90E-05 | 5,20E-05 | 0 | 0,99701 | 0,000184 | 0 | 0,000184 | 0 | 5,20E-05 | 0 | 0 | 5,20E-05 | 0 | 0 | 0 | 7,90E-05 | 0 | 0 | 0,000865 | 0 | 5,20E-05 | 0 | 5,20E-05 | 5,20E-05 | 0,000551 |
| **9** | 0,000874 | 0,000874 | 0,001661 | 0,000175 | 0,000874 | 0,000175 | 0,000437 | 0,000787 | 0,988372 | 0 | 0,000612 | 0 | 0,003147 | 0 | 0 | 0 | 0 | 0 | 0 | 0 | 0 | 0 | 0,001049 | 0 | 0,000175 | 0 | 0 | 0 | 0,000787 |
| **11** | 0 | 0,000427 | 0 | 0 | 0,000142 | 0 | 0 | 0 | 0 | 0,998151 | 0 | 0 | 0 | 0 | 0 | 0 | 0 | 0 | 0 | 0 | 0 | 0 | 0,000569 | 0 | 0 | 0 | 0 | 0 | 0,000711 |
| **12** | 2,50E-05 | 2,50E-05 | 0 | 0 | 0 | 0 | 0 | 0,000175 | 0,00015 | 0 | 0,996926 | 2,50E-05 | 0 | 0 | 0 | 0 | 0 | 0 | 0 | 0 | 0 | 0 | 0,001849 | 0 | 7,50E-05 | 0 | 0 | 2,50E-05 | 0,000725 |
| **13** | 0 | 0 | 0,0625 | 0 | 0 | 0 | 0 | 0 | 0,0625 | 0 | 0,0625 | 0,8125 | 0 | 0 | 0 | 0 | 0 | 0 | 0 | 0 | 0 | 0 | 0 | 0 | 0 | 0 | 0 | 0 | 0 |
| **14** | 2,30E-05 | 2,30E-05 | 3,40E-05 | 0 | 5,70E-05 | 0 | 1,10E-05 | 4,60E-05 | 0,000402 | 0 | 1,10E-05 | 0 | 0,998794 | 0 | 0 | 0 | 0 | 0 | 0 | 0 | 0 | 0 | 5,70E-05 | 0 | 1,10E-05 | 0 | 1,10E-05 | 0 | 0,000517 |
| **16** | 0 | 0 | 0 | 0 | 0 | 0 | 0 | 0 | 0 | 0 | 0 | 0 | 0 | 0 | 0 | 0 | 0 | 0 | 0 | 0 | 0 | 0 | 0 | 0 | 0 | 0 | 0 | 0 | 0 |
| **17** | 0 | 0 | 0 | 0 | 0 | 0 | 0 | 0 | 0 | 0 | 0 | 0 | 0 | 0 | 0 | 0 | 0 | 0 | 0 | 0 | 0 | 0 | 0 | 0 | 0 | 0 | 0 | 0 | 0 |
| **18** | 0 | 0,020833 | 0,010417 | 0 | 0 | 0 | 0 | 0,010417 | 0 | 0 | 0 | 0 | 0,010417 | 0 | 0 | 0,927083 | 0 | 0 | 0 | 0 | 0 | 0,010417 | 0 | 0 | 0 | 0 | 0,010417 | 0 | 0 |
| **20** | 0 | 0 | 0 | 0 | 0 | 0 | 0 | 0 | 0 | 0 | 0 | 0 | 0 | 0 | 0 | 0 | 0 | 0 | 0 | 0 | 0 | 0 | 0 | 0 | 0 | 0 | 0 | 0 | 0 |
| **21** | 0 | 0 | 0 | 0 | 0 | 0 | 0 | 0 | 0 | 0 | 0 | 0 | 0 | 0 | 0 | 0 | 0 | 0 | 0 | 0 | 0 | 0 | 0 | 0 | 0 | 0 | 0 | 0 | 0 |
| **22** | 0 | 0 | 0 | 0 | 0 | 0 | 0 | 0 | 0 | 0 | 0 | 0 | 0 | 0 | 0 | 0 | 0 | 0 | 0 | 0 | 0 | 0 | 0 | 0 | 0 | 0 | 0 | 0 | 0 |
| **23** | 0 | 0 | 0 | 0 | 0 | 0 | 0 | 0,040541 | 0 | 0 | 0 | 0 | 0 | 0 | 0 | 0 | 0 | 0 | 0 | 0,959459 | 0 | 0 | 0 | 0 | 0 | 0 | 0 | 0 | 0 |
| **24** | 0 | 0 | 0 | 0 | 0 | 0 | 0 | 0 | 0 | 0 | 0 | 0 | 0 | 0 | 0 | 0 | 0 | 0 | 0 | 0 | 0 | 0 | 0 | 0 | 0 | 0 | 0 | 0 | 0 |
| **25** | 0 | 0 | 0 | 0 | 0,013889 | 0 | 0 | 0 | 0 | 0 | 0 | 0 | 0 | 0 | 0 | 0,013889 | 0 | 0 | 0 | 0 | 0 | 0,972222 | 0 | 0 | 0 | 0 | 0 | 0 | 0 |
| **26** | 0,005892 | 0,012319 | 0,011784 | 0,003214 | 0,004017 | 0,000803 | 0,00241 | 0,008034 | 0,00616 | 0,001339 | 0,019014 | 0 | 0,001875 | 0 | 0 | 0 | 0 | 0 | 0 | 0 | 0 | 0 | 0,905731 | 0 | 0,000268 | 0,001339 | 0,015801 | 0 | 0 |
| **27** | 0 | 0 | 0 | 0 | 0 | 0 | 0 | 0 | 0 | 0 | 0 | 0 | 0 | 0 | 0 | 0 | 0 | 0 | 0 | 0 | 0 | 0 | 0 | 0 | 0 | 0 | 0 | 0 | 0 |
| **28** | 0,014577 | 0,020408 | 0,011662 | 0 | 0 | 0,002915 | 0 | 0,008746 | 0 | 0 | 0,011662 | 0 | 0,002915 | 0 | 0 | 0 | 0 | 0 | 0 | 0 | 0 | 0 | 0 | 0 | 0,915452 | 0 | 0,005831 | 0,005831 | 0 |
| **29** | 0 | 0 | 0 | 0 | 0 | 0 | 0 | 0 | 0 | 0 | 0 | 0 | 0 | 0 | 0 | 0 | 0 | 0 | 0 | 0 | 0 | 0 | 0,001572 | 0 | 0 | 0,997799 | 0 | 0 | 0,000629 |
| **30** | 0,002532 | 0,003271 | 0,002849 | 0 | 0,000739 | 0,000211 | 0,000317 | 0,002427 | 0,001161 | 0,000106 | 0,000317 | 0 | 0,000211 | 0 | 0 | 0 | 0 | 0 | 0 | 0 | 0 | 0 | 0,006753 | 0 | 0,000422 | 0 | 0,978685 | 0 | 0 |
| **31** | 0,000201 | 0 | 0 | 0 | 0 | 0 | 0 | 1,00E-04 | 0 | 0 | 0 | 0 | 0 | 0 | 0 | 0 | 0 | 0 | 0 | 0 | 0 | 0 | 0 | 0 | 1,00E-04 | 0 | 0 | 0,999197 | 0,000402 |
| **out** | 0 | 0 | 0 | 0 | 0 | 0 | 0 | 0 | 0 | 0 | 0 | 0 | 0 | 0 | 0 | 0 | 0 | 0 | 0 | 0 | 0 | 0 | 0 | 0 | 0 | 0 | 0 | 0 | 1 |

**Table S4.** Transition matrix for patients hospitalized in the internal medicine department

**Table S5.** Ward-specific probabilities of undergoing each of the procedures in the surgery department

|  | **surgery** | **intravenous** | **sutures** | **blood_transfusion** | **blood_sample** | **injection** | **endoscopy** | **gastric_lavage** | **cardiac_catheter** | **dialysis** | **wound_dressing** | **bloodglucose** | **endotrachealintu** | **drainagecatheter** | **other_invproc** | **no_proc** |
| --- | --- | --- | --- | --- | --- | --- | --- | --- | --- | --- | --- | --- | --- | --- | --- | --- |
| **1** | 0 | 0 | 0 | 0 | 0 | 0 | 0 | 0 | 0 | 0 | 0 | 0 | 0 | 0 | 0 | 1 |
| **2** | 0 | 0 | 0 | 0 | 0 | 0 | 0 | 0 | 0 | 0 | 0 | 0 | 0 | 0 | 0 | 1 |
| **3** | 0 | 0 | 0 | 0 | 0 | 0 | 0 | 0 | 0 | 0 | 0 | 0 | 0 | 0 | 0 | 1 |
| **4** | 0 | 0 | 0 | 0 | 0 | 0 | 0 | 0 | 0 | 0 | 0 | 0 | 0 | 0 | 0 | 1 |
| **5** | 0 | 0 | 0 | 0 | 0 | 0 | 0 | 0 | 0 | 0 | 0 | 0 | 0 | 0 | 0 | 1 |
| **6** | 0 | 0 | 0 | 0 | 0 | 0 | 0 | 0 | 0 | 0 | 0 | 0 | 0 | 0 | 0 | 1 |
| **7** | 0 | 0 | 0 | 0 | 0 | 0 | 0 | 0 | 0 | 0 | 0 | 0 | 0 | 0 | 0 | 1 |
| **8** | 0 | 0 | 0 | 0 | 0 | 0 | 0 | 0 | 0 | 0 | 0 | 0 | 0 | 0 | 0 | 1 |
| **9** | 0 | 0 | 0 | 0 | 0 | 0 | 0 | 0 | 0 | 0 | 0 | 0 | 0 | 0 | 0 | 1 |
| **11** | 0 | 0 | 0 | 0 | 0 | 0 | 0 | 0 | 0 | 0 | 0 | 0 | 0 | 0 | 0 | 1 |
| **12** | 0 | 0 | 0 | 0 | 0 | 0 | 0 | 0 | 0 | 0 | 0 | 0 | 0 | 0 | 0 | 1 |
| **13** | 0 | 0 | 0 | 0 | 0 | 0 | 0 | 0 | 0 | 0 | 0 | 0 | 0 | 0 | 0 | 1 |
| **14** | 0 | 0 | 0 | 0 | 0 | 0 | 0 | 0 | 0 | 0 | 0 | 0 | 0 | 0 | 0 | 1 |
| **16** | 0 | 0 | 0 | 0 | 0,004766 | 0,000636 | 0 | 0 | 0 | 0 | 0 | 0,010168 | 0 | 0 | 0,000636 | 0,983794 |
| **17** | 0 | 0,000289 | 0 | 0 | 0,001235 | 0,005381 | 2,30E-05 | 0 | 0 | 0 | 0,000208 | 0,000312 | 0 | 0 | 0 | 0,992552 |
| **18** | 0 | 0,000714 | 9,00E-06 | 1,70E-05 | 0,001239 | 0,001093 | 0 | 0 | 0 | 0 | 0,000439 | 0,000404 | 0 | 0 | 0 | 0,996086 |
| **20** | 0,000107 | 0,00075 | 0 | 0 | 0,000536 | 0,001929 | 0 | 0 | 0 | 0 | 0 | 0,00225 | 0 | 0 | 0 | 0,994428 |
| **21** | 0 | 0,000454 | 0 | 0 | 0,001362 | 0,000681 | 0 | 0 | 0 | 0 | 0 | 0 | 0 | 0 | 0 | 0,997503 |
| **22** | 0 | 0 | 0 | 0 | 0 | 0 | 0 | 0 | 0 | 0 | 0 | 0 | 0 | 0 | 0 | 1 |
| **23** | 0 | 0 | 0 | 0 | 0 | 0 | 0 | 0 | 0 | 0 | 0 | 0 | 0 | 0 | 0 | 1 |
| **24** | 0 | 0,001161 | 0 | 0 | 0,001347 | 0,001254 | 0 | 0 | 0 | 0 | 0,001114 | 4,60E-05 | 0 | 0 | 0 | 0,995078 |
| **25** | 0,027213 | 0,02567 | 0,022444 | 0,000421 | 0 | 0,002946 | 0,00014 | 0 | 0 | 0 | 0,000701 | 0,00014 | 0,023145 | 0,006453 | 0,005471 | 0,885257 |
| **26** | 0 | 0,001138 | 0 | 0 | 0 | 0 | 0 | 0 | 0 | 0 | 0 | 0 | 0 | 0 | 0 | 0,998862 |
| **27** | 0 | 0 | 0 | 0 | 0,159427 | 0 | 0 | 0 | 0 | 0 | 0 | 0 | 0,002311 | 0 | 0 | 0,838262 |
| **28** | 0 | 0 | 0 | 0 | 0 | 0 | 0,022989 | 0 | 0 | 0 | 0 | 0 | 0 | 0 | 0 | 0,977011 |
| **29** | 0 | 0 | 0 | 0 | 0 | 0 | 0 | 0 | 0 | 0 | 0 | 0 | 0 | 0 | 0 | 1 |
| **30** | 0 | 0,015219 | 0,000231 | 0 | 0,023289 | 0,001384 | 0 | 0 | 0 | 0 | 0,000692 | 0,000231 | 0 | 0 | 0,000461 | 0,958495 |
| **31** | 0 | 0 | 0 | 0 | 0 | 0 | 0 | 0 | 0 | 0 | 0 | 0 | 0 | 0 | 0 | 1 |

**Table S6.** Ward-specific probabilities of undergoing each of the procedures in the internal medicine department

|  | **surgery** | **intravenous** | **sutures** | **blood_transfusion** | **blood_sample** | **injection** | **endoscopy** | **gastric_lavage** | **cardiac_catheter** | **dialysis** | **wound_dressing** | **bloodglucose** | **endotrachealintu** | **drainagecatheter** | **other_invproc** | **no_proc** |
| --- | --- | --- | --- | --- | --- | --- | --- | --- | --- | --- | --- | --- | --- | --- | --- | --- |
| **1** | 0 | 0,000671 | 0 | 0,000307 | 0,002989 | 0,000838 | 5,60E-05 | 8,40E-05 | 0 | 0 | 0 | 0,00109 | 0 | 0 | 0,000643 | 0,993323 |
| **2** | 0 | 0,000825 | 0 | 0,000474 | 0,003319 | 0,000577 | 0 | 0 | 0 | 0 | 0 | 0,000495 | 0 | 0 | 0,000124 | 0,994186 |
| **3** | 0 | 0,000814 | 0 | 0,000352 | 0,003629 | 0,001342 | 0 | 0 | 0 | 0 | 0 | 0,001254 | 0 | 0 | 4,40E-05 | 0,992565 |
| **4** | 0 | 0,001027 | 0 | 0,000486 | 0,003404 | 0,001243 | 0 | 0 | 0 | 0 | 0 | 0 | 0 | 0 | 0,000216 | 0,993625 |
| **5** | 0 | 0,001335 | 0 | 0,000103 | 0,003593 | 0,001386 | 0 | 0 | 0 | 0 | 0 | 0,001437 | 0 | 0 | 0 | 0,992147 |
| **6** | 0 | 0,000338 | 0 | 0,000169 | 0,002618 | 0,001267 | 0 | 0 | 0 | 0 | 0 | 0,0038 | 0 | 0 | 0,000507 | 0,991301 |
| **7** | 0 | 0,000792 | 0 | 0 | 0,003961 | 0,000226 | 0 | 0 | 0 | 0 | 0 | 0,000622 | 0 | 0 | 0,00017 | 0,994228 |
| **8** | 0 | 0,000839 | 5,20E-05 | 0,000446 | 0,002937 | 0,002727 | 0 | 0 | 2,60E-05 | 0,001049 | 0 | 0,001914 | 0 | 2,60E-05 | 0,000367 | 0,989615 |
| **9** | 0 | 0,000525 | 8,70E-05 | 0 | 0,002798 | 0,006557 | 0 | 0 | 0,000525 | 0 | 0 | 0,003934 | 0 | 0 | 0,000437 | 0,985137 |
| **11** | 0 | 0,001422 | 0 | 0 | 0,002133 | 0,003981 | 0,000142 | 0 | 0 | 0 | 0 | 0,003981 | 0 | 0 | 0 | 0,98834 |
| **12** | 0 | 9,00E-04 | 0 | 5,00E-04 | 0,002699 | 0,002024 | 0 | 5,00E-05 | 0 | 0 | 0 | 0,001524 | 0 | 0 | 0,000175 | 0,992129 |
| **13** | 0 | 0 | 0 | 0 | 0 | 0 | 0 | 0 | 0 | 0 | 0 | 0 | 0 | 0 | 0 | 1 |
| **14** | 0,000425 | 0,000161 | 0 | 0 | 0,000769 | 0,001183 | 0 | 0 | 0 | 0 | 0 | 7,00E-04 | 3,40E-05 | 0 | 0 | 0,996728 |
| **16** | 0 | 0 | 0 | 0 | 0 | 0 | 0 | 0 | 0 | 0 | 0 | 0 | 0 | 0 | 0 | 1 |
| **17** | 0 | 0 | 0 | 0 | 0 | 0 | 0 | 0 | 0 | 0 | 0 | 0 | 0 | 0 | 0 | 1 |
| **18** | 0 | 0 | 0 | 0 | 0 | 0 | 0 | 0 | 0 | 0 | 0 | 0 | 0 | 0 | 0 | 1 |
| **20** | 0 | 0 | 0 | 0 | 0 | 0 | 0 | 0 | 0 | 0 | 0 | 0 | 0 | 0 | 0 | 1 |
| **21** | 0 | 0 | 0 | 0 | 0 | 0 | 0 | 0 | 0 | 0 | 0 | 0 | 0 | 0 | 0 | 1 |
| **22** | 0 | 0 | 0 | 0 | 0 | 0 | 0 | 0 | 0 | 0 | 0 | 0 | 0 | 0 | 0 | 1 |
| **23** | 0 | 0 | 0 | 0 | 0 | 0 | 0 | 0 | 0 | 0 | 0 | 0 | 0 | 0 | 0 | 1 |
| **24** | 0 | 0 | 0 | 0 | 0 | 0 | 0 | 0 | 0 | 0 | 0 | 0 | 0 | 0 | 0 | 1 |
| **25** | 0,027778 | 0 | 0 | 0,013889 | 0 | 0,027778 | 0 | 0 | 0 | 0 | 0 | 0 | 0 | 0 | 0 | 0,930556 |
| **26** | 0,001339 | 0,001071 | 0 | 0,000268 | 0 | 0,00241 | 0,000268 | 0 | 0 | 0 | 0 | 0 | 0 | 0 | 0,004016 | 0,990628 |
| **27** | 0 | 0 | 0 | 0 | 0 | 0 | 0 | 0 | 0 | 0 | 0 | 0 | 0 | 0 | 0 | 1 |
| **28** | 0 | 0,002915 | 0 | 0 | 0,002915 | 0,002915 | 0,052478 | 0,002915 | 0 | 0 | 0 | 0 | 0 | 0 | 0 | 0,93586 |
| **29** | 0 | 0,000314 | 0 | 0 | 0,001572 | 0,000314 | 0 | 0 | 0 | 0 | 0 | 0 | 0 | 0 | 0 | 0,997799 |
| **30** | 0,000106 | 0,012239 | 0 | 0,001266 | 0,014771 | 0,001794 | 0 | 0,001372 | 0 | 0 | 0 | 0,003798 | 0 | 0 | 0,001583 | 0,963072 |
| **31** | 0 | 0,000502 | 1,00E-04 | 0,001305 | 0,003213 | 0,00241 | 0 | 0,001305 | 0 | 1,00E-04 | 0,001104 | 0,011245 | 0,000502 | 0 | 0,001506 | 0,976707 |

| **Ward**  **Device** | **1** | **2** | **3** | **4** | **5** | **6** | **7** | **8** | **9** | **11** | **12** | **13** | **14** | **16** | **17** | **18** | **20** | **21** | **22** | **23** | **24** | **25** | **26** | **27** | **28** | **29** | **30** | **31** |
| --- | --- | --- | --- | --- | --- | --- | --- | --- | --- | --- | --- | --- | --- | --- | --- | --- | --- | --- | --- | --- | --- | --- | --- | --- | --- | --- | --- | --- |
| **Syringes** | 1155263 | 1654356 | 1984501 | 708925 | 923282 | 484672 | 577928 | 1884121 | 880828 | 397425 | 1596235 | 0 | 1775710 | 81079 | 2858066 | 1346126 | 112612 | 46574 | 0 | 0 | 266490 | 1012436 | 119147 | 357612 | 306 | 48462 | 39879 | 419600 |
| **IV Set** | 21171 | 40436 | 33712 | 15993 | 19258 | 4664 | 7657 | 56117 | 7030 | 6760 | 33219 | 0 | 33252 | 0 | 58692 | 205191 | 17417 | 5204 | 0 | 0 | 59330 | 845891 | 11308 | 0 | 162 | 576 | 30441 | 10147 |
| **IV Cannula** | 13550 | 25878 | 21571 | 10237 | 12327 | 2984 | 4900 | 35914 | 4498 | 4325 | 21261 | 0 | 21278 | 0 | 28756 | 100547 | 8537 | 2548 | 0 | 0 | 29069 | 415002 | 6613 | 0 | 162 | 371 | 30441 | 6497 |
| **Scalpel** | 0 | 0 | 0 | 0 | 0 | 0 | 0 | 0 | 0 | 0 | 0 | 0 | 17263880 | 0 | 0 | 0 | 55 | 0 | 0 | 0 | 0 | 39434 | 2149057 | 0 | 0 | 0 | 144 | 0 |
| **Lancet** | 15249 | 9251 | 23820 | 0 | 12718 | 22324 | 3974 | 30616 | 17948 | 12712 | 24539 | 0 | 23393 | 685564 | 615017 | 1163877 | 449300 | 0 | 0 | 0 | 20551 | 25776 | 0 | 0 | 0 | 0 | 6046 | 39707 |
| **Surgical needles & suture kits** | 604358 | 813480 | 810317 | 312197 | 369568 | 214978 | 316054 | 619479 | 179875 | 78270 | 546694 | 0 | 504489 | 20409490 | 159796014 | 237981903 | 7371332 | 7761866 | 0 | 0 | 62398459 | 444178020 | 93270 | 88643180 | 160 | 21811 | 39652 | 253788 |
| **Endotracheal tube** | 0 | 0 | 0 | 0 | 0 | 0 | 0 | 0 | 0 | 0 | 0 | 0 | 4331 | 0 | 0 | 0 | 15 | 0 | 0 | 0 | 0 | 4843 | 498 | 16 | 0 | 0 | 144 | 489 |
| **Drainage catheter** | 0 | 0 | 0 | 0 | 0 | 0 | 0 | 145 | 0 | 0 | 0 | 0 | 0 | 0 | 0 | 0 | 0 | 0 | 0 | 0 | 0 | 6867 | 0 | 0 | 0 | 0 | 0 | 0 |
| **Gastric lavage tube** | 405 | 0 | 0 | 0 | 0 | 0 | 0 | 0 | 0 | 0 | 334 | 0 | 0 | 0 | 0 | 0 | 0 | 0 | 0 | 0 | 0 | 0 | 0 | 0 | 149 | 0 | 2086 | 2316 |
| **Endoscope** | 4096 | 0 | 0 | 0 | 0 | 0 | 0 | 0 | 0 | 2507 | 0 | 0 | 0 | 0 | 8396 | 0 | 0 | 0 | 0 | 0 | 0 | 4688 | 2289 | 0 | 2978 | 0 | 0 | 0 |

**Table S7.** Yearly initial quantity of new devices in each ward for the high-resource setting

**Table S8.** Yearly initial quantity of previously used devices in each ward for the high-resource setting

| **Ward**  **Device** | **1** | **2** | **3** | **4** | **5** | **6** | **7** | **8** | **9** | **11** | **12** | **13** | **14** | **16** | **17** | **18** | **20** | **21** | **22** | **23** | **24** | **25** | **26** | **27** | **28** | **29** | **30** | **31** |
| --- | --- | --- | --- | --- | --- | --- | --- | --- | --- | --- | --- | --- | --- | --- | --- | --- | --- | --- | --- | --- | --- | --- | --- | --- | --- | --- | --- | --- |
| **Syringes** | 0 | 0 | 0 | 0 | 0 | 0 | 0 | 0 | 0 | 0 | 0 | 0 | 0 | 0 | 0 | 0 | 0 | 0 | 0 | 0 | 0 | 0 | 0 | 0 | 0 | 0 | 0 | 0 |
| **IV Set** | 0 | 0 | 0 | 0 | 0 | 0 | 0 | 0 | 0 | 0 | 0 | 0 | 0 | 0 | 0 | 0 | 0 | 0 | 0 | 0 | 0 | 0 | 0 | 0 | 0 | 0 | 0 | 0 |
| **IV Cannula** | 0 | 0 | 0 | 0 | 0 | 0 | 0 | 0 | 0 | 0 | 0 | 0 | 0 | 0 | 0 | 0 | 0 | 0 | 0 | 0 | 0 | 0 | 0 | 0 | 0 | 0 | 0 | 0 |
| **Scalpel** | 0 | 0 | 0 | 0 | 0 | 0 | 0 | 0 | 0 | 0 | 0 | 0 | 0 | 0 | 0 | 0 | 30 | 0 | 0 | 0 | 0 | 0 | 0 | 0 | 0 | 0 | 0 | 0 |
| **Lancet** | 0 | 0 | 0 | 0 | 0 | 0 | 0 | 0 | 0 | 0 | 0 | 0 | 0 | 0 | 0 | 0 | 0 | 0 | 0 | 0 | 0 | 0 | 0 | 0 | 0 | 0 | 0 | 0 |
| **Surgical needles & suture kits** | 0 | 0 | 0 | 0 | 0 | 0 | 0 | 0 | 0 | 0 | 0 | 0 | 0 | 0 | 0 | 0 | 0 | 0 | 0 | 0 | 0 | 0 | 0 | 0 | 0 | 0 | 0 | 0 |
| **Endotracheal tube** | 0 | 0 | 0 | 0 | 0 | 0 | 0 | 0 | 0 | 0 | 0 | 0 | 861 | 0 | 0 | 0 | 29 | 0 | 0 | 0 | 0 | 11079 | 151 | 28 | 0 | 0 | 0 | 160 |
| **Drainage catheter** | 0 | 0 | 0 | 0 | 0 | 0 | 0 | 0 | 0 | 0 | 0 | 0 | 0 | 0 | 0 | 0 | 0 | 0 | 0 | 0 | 0 | 0 | 0 | 0 | 0 | 0 | 0 | 0 |
| **Gastric lavage tube** | 0 | 0 | 0 | 0 | 0 | 0 | 0 | 0 | 0 | 0 | 0 | 0 | 0 | 0 | 0 | 0 | 0 | 0 | 0 | 0 | 0 | 0 | 0 | 0 | 0 | 0 | 0 | 0 |
| **Endoscope** | 0 | 0 | 0 | 0 | 0 | 0 | 0 | 0 | 0 | 0 | 0 | 0 | 0 | 0 | 0 | 0 | 0 | 0 | 0 | 0 | 0 | 0 | 0 | 0 | 0 | 0 | 0 | 0 |

**Table S9.** Yearly initial quantity of new devices in each ward for the low-resource setting

| **Ward**  **Device** | **1** | **2** | **3** | **4** | **5** | **6** | **7** | **8** | **9** | **11** | **12** | **13** | **14** | **16** | **17** | **18** | **20** | **21** | **22** | **23** | **24** | **25** | **26** | **27** | **28** | **29** | **30** | **31** |
| --- | --- | --- | --- | --- | --- | --- | --- | --- | --- | --- | --- | --- | --- | --- | --- | --- | --- | --- | --- | --- | --- | --- | --- | --- | --- | --- | --- | --- |
| **Syringes** | 288816 | 413589 | 496125 | 177231 | 230821 | 121168 | 144482 | 471030 | 220207 | 99356 | 399059 | 0 | 443928 | 20270 | 714516 | 336531 | 28153 | 11644 | 0 | 0 | 66623 | 253109 | 29787 | 89403 | 76 | 12116 | 9970 | 104900 |
| **IV Set** | 5293 | 10109 | 8428 | 3998 | 4814 | 1166 | 1914 | 14029 | 1757 | 1690 | 8305 | 0 | 8313 | 0 | 14673 | 51298 | 4354 | 1301 | 0 | 0 | 14833 | 211473 | 2827 | 0 | 40 | 144 | 7610 | 2537 |
| **IV Cannula** | 3387 | 6470 | 5393 | 2559 | 3082 | 746 | 1225 | 8979 | 1124 | 1081 | 5315 | 0 | 5320 | 0 | 7189 | 25137 | 2134 | 637 | 0 | 0 | 7267 | 103751 | 1653 | 0 | 40 | 93 | 7610 | 1624 |
| **Scalpel** | 0 | 0 | 0 | 0 | 0 | 0 | 0 | 0 | 0 | 0 | 0 | 0 | 4315970 | 0 | 0 | 0 | 14 | 0 | 0 | 0 | 0 | 9859 | 537264 | 0 | 0 | 0 | 36 | 0 |
| **Lancet** | 3812 | 2313 | 5955 | 0 | 3180 | 5581 | 994 | 7654 | 4487 | 3178 | 6135 | 0 | 5848 | 171391 | 153754 | 290969 | 112325 | 0 | 0 | 0 | 5138 | 6444 | 0 | 0 | 0 | 0 | 1512 | 9927 |
| **Surgical needles & suture kits** | 151090 | 203370 | 202579 | 78049 | 92392 | 53744 | 79013 | 154870 | 44969 | 19567 | 136674 | 0 | 126122 | 5102373 | 39949004 | 59495476 | 1842833 | 1940466 | 0 | 0 | 15599615 | 111044505 | 23318 | 22160795 | 40 | 5453 | 9913 | 63447 |
| **Endotracheal tube** | 0 | 0 | 0 | 0 | 0 | 0 | 0 | 0 | 0 | 0 | 0 | 0 | 1083 | 0 | 0 | 0 | 4 | 0 | 0 | 0 | 0 | 1211 | 125 | 4 | 0 | 0 | 36 | 122 |
| **Drainage catheter** | 0 | 0 | 0 | 0 | 0 | 0 | 0 | 36 | 0 | 0 | 0 | 0 | 0 | 0 | 0 | 0 | 0 | 0 | 0 | 0 | 0 | 1717 | 0 | 0 | 0 | 0 | 0 | 0 |
| **Gastric lavage tube** | 101 | 0 | 0 | 0 | 0 | 0 | 0 | 0 | 0 | 0 | 84 | 0 | 0 | 0 | 0 | 0 | 0 | 0 | 0 | 0 | 0 | 0 | 0 | 0 | 37 | 0 | 522 | 579 |
| **Endoscope** | 1024 | 0 | 0 | 0 | 0 | 0 | 0 | 0 | 0 | 627 | 0 | 0 | 0 | 0 | 2099 | 0 | 0 | 0 | 0 | 0 | 0 | 1172 | 572 | 0 | 744 | 0 | 0 | 0 |

**Table S10.** Yearly initial quantity of previously used devices in each ward for the low-resource setting

| **Ward**  **Device** | **1** | **2** | **3** | **4** | **5** | **6** | **7** | **8** | **9** | **11** | **12** | **13** | **14** | **16** | **17** | **18** | **20** | **21** | **22** | **23** | **24** | **25** | **26** | **27** | **28** | **29** | **30** | **31** |
| --- | --- | --- | --- | --- | --- | --- | --- | --- | --- | --- | --- | --- | --- | --- | --- | --- | --- | --- | --- | --- | --- | --- | --- | --- | --- | --- | --- | --- |
| **Syringes** | 0 | 0 | 0 | 0 | 0 | 0 | 0 | 0 | 0 | 0 | 0 | 0 | 0 | 0 | 0 | 0 | 0 | 0 | 0 | 0 | 0 | 0 | 0 | 0 | 76 | 0 | 9970 | 0 |
| **IV Set** | 188 | 0 | 0 | 386 | 0 | 0 | 278 | 63 | 122 | 0 | 463 | 0 | 0 | 0 | 0 | 0 | 0 | 0 | 0 | 0 | 0 | 0 | 0 | 0 | 40 | 12 | 7610 | 438 |
| **IV Cannula** | 997 | 1422 | 1246 | 948 | 426 | 5 | 528 | 2295 | 379 | 171 | 1699 | 0 | 1068 | 0 | 0 | 0 | 0 | 0 | 0 | 0 | 0 | 0 | 196 | 0 | 40 | 33 | 7610 | 756 |
| **Scalpel** | 0 | 0 | 0 | 0 | 0 | 0 | 0 | 0 | 0 | 0 | 0 | 0 | 0 | 0 | 0 | 0 | 29 | 0 | 0 | 0 | 0 | 6871 | 0 | 0 | 0 | 0 | 36 | 0 |
| **Lancet** | 1196 | 769 | 1365 | 0 | 416 | 198 | 419 | 1721 | 1292 | 418 | 1699 | 0 | 1986 | 0 | 0 | 0 | 0 | 0 | 0 | 0 | 0 | 0 | 0 | 0 | 0 | 0 | 1512 | 4457 |
| **Surgical needles & suture kits** | 0 | 0 | 0 | 0 | 0 | 0 | 0 | 0 | 0 | 0 | 0 | 0 | 0 | 0 | 0 | 0 | 0 | 0 | 0 | 0 | 0 | 0 | 0 | 0 | 40 | 0 | 9913 | 0 |
| **Endotracheal tube** | 0 | 0 | 0 | 0 | 0 | 0 | 0 | 0 | 0 | 0 | 0 | 0 | 1513 | 0 | 0 | 0 | 18 | 0 | 0 | 0 | 0 | 6750 | 200 | 18 | 0 | 0 | 36 | 202 |
| **Drainage catheter** | 0 | 0 | 0 | 0 | 0 | 0 | 0 | 36 | 0 | 0 | 0 | 0 | 0 | 0 | 0 | 0 | 0 | 0 | 0 | 0 | 0 | 1717 | 0 | 0 | 0 | 0 | 0 | 0 |
| **Gastric lavage tube** | 101 | 0 | 0 | 0 | 0 | 0 | 0 | 0 | 0 | 0 | 84 | 0 | 0 | 0 | 0 | 0 | 0 | 0 | 0 | 0 | 0 | 0 | 0 | 0 | 37 | 0 | 522 | 579 |
| **Endoscope** | 0 | 0 | 0 | 0 | 0 | 0 | 0 | 0 | 0 | 0 | 0 | 0 | 0 | 0 | 0 | 0 | 0 | 0 | 0 | 0 | 0 | 0 | 0 | 0 | 744 | 0 | 0 | 0 |

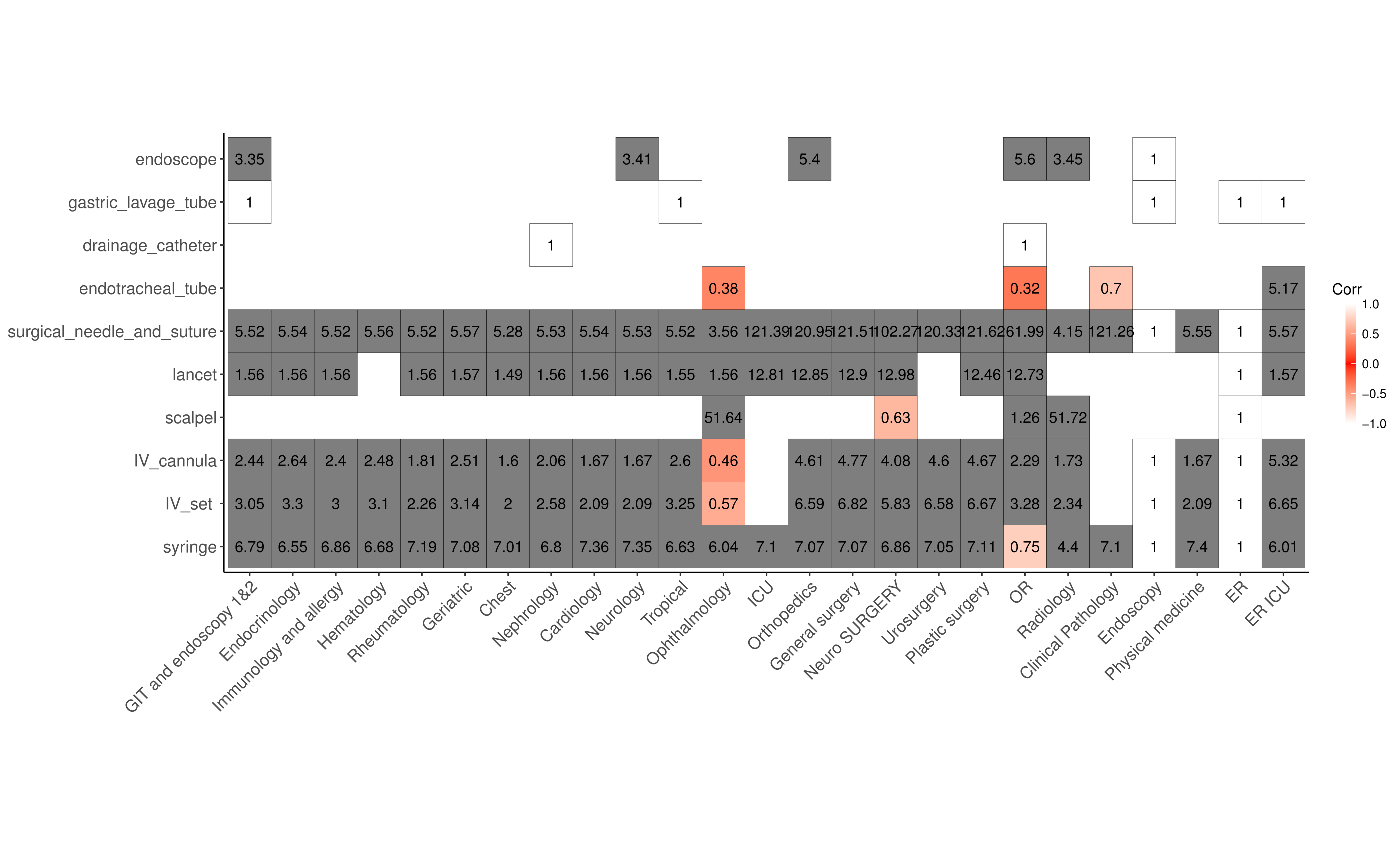

**Figure S1.** Use of devices over available quantity of devices for each type in each ward in the high-resource hospital. There is insufficient devices when the value < 1.

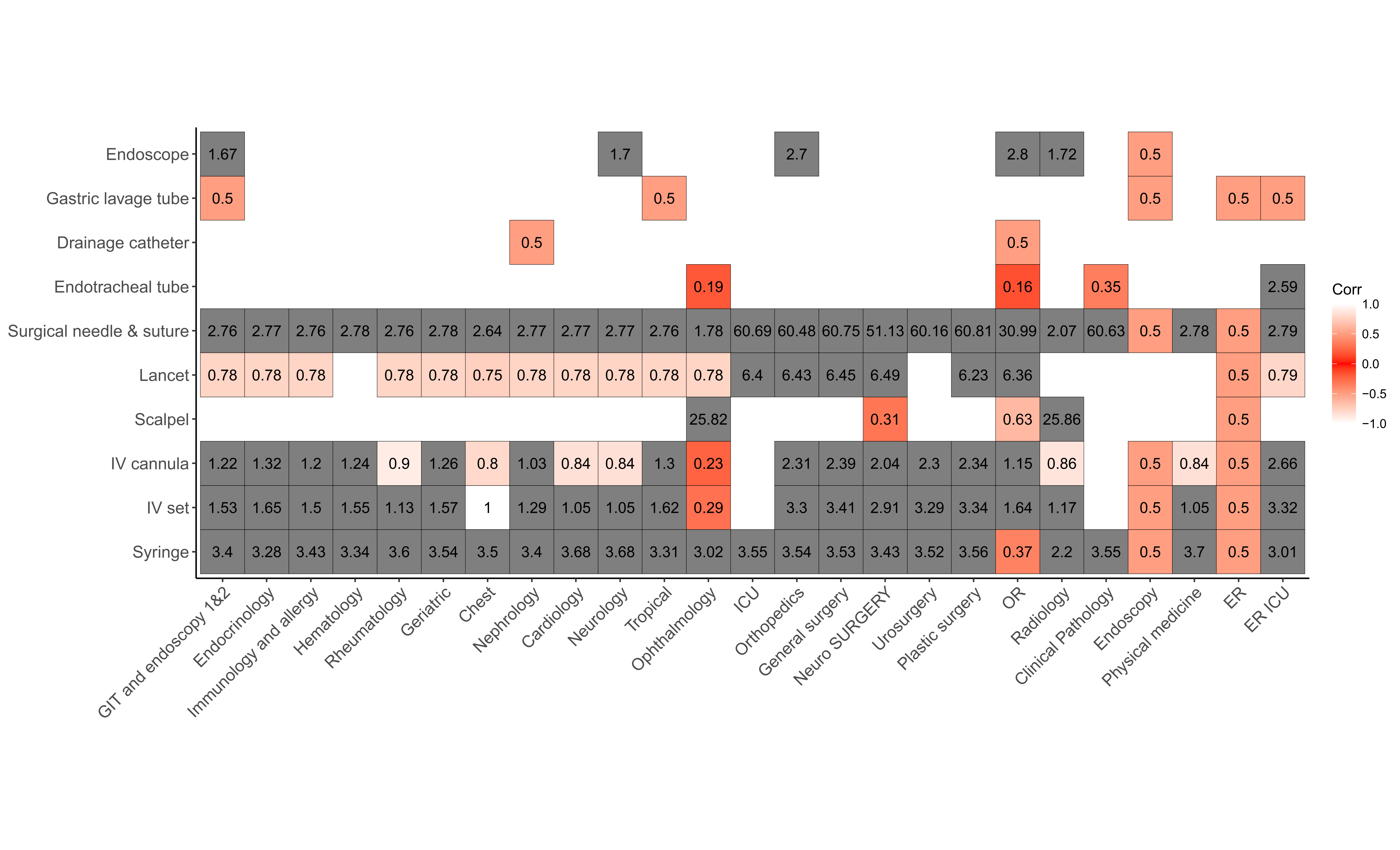

**Figure S2**. Use of devices over available quantity of devices for each type in each ward in the low-resource hospital. There is insufficient devices when the value < 1.

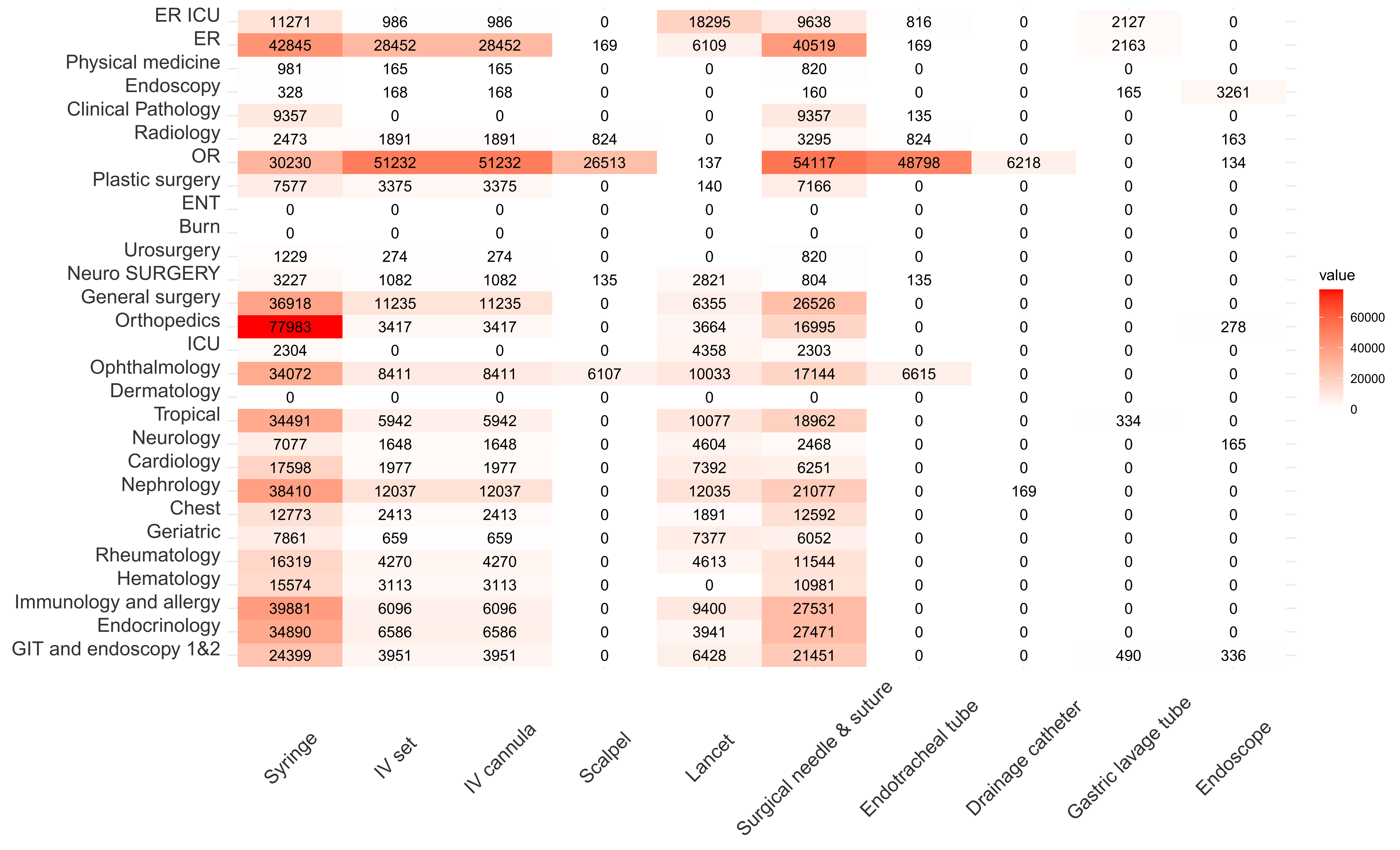

**Figure S3.** Yearly equipment uses in each ward.

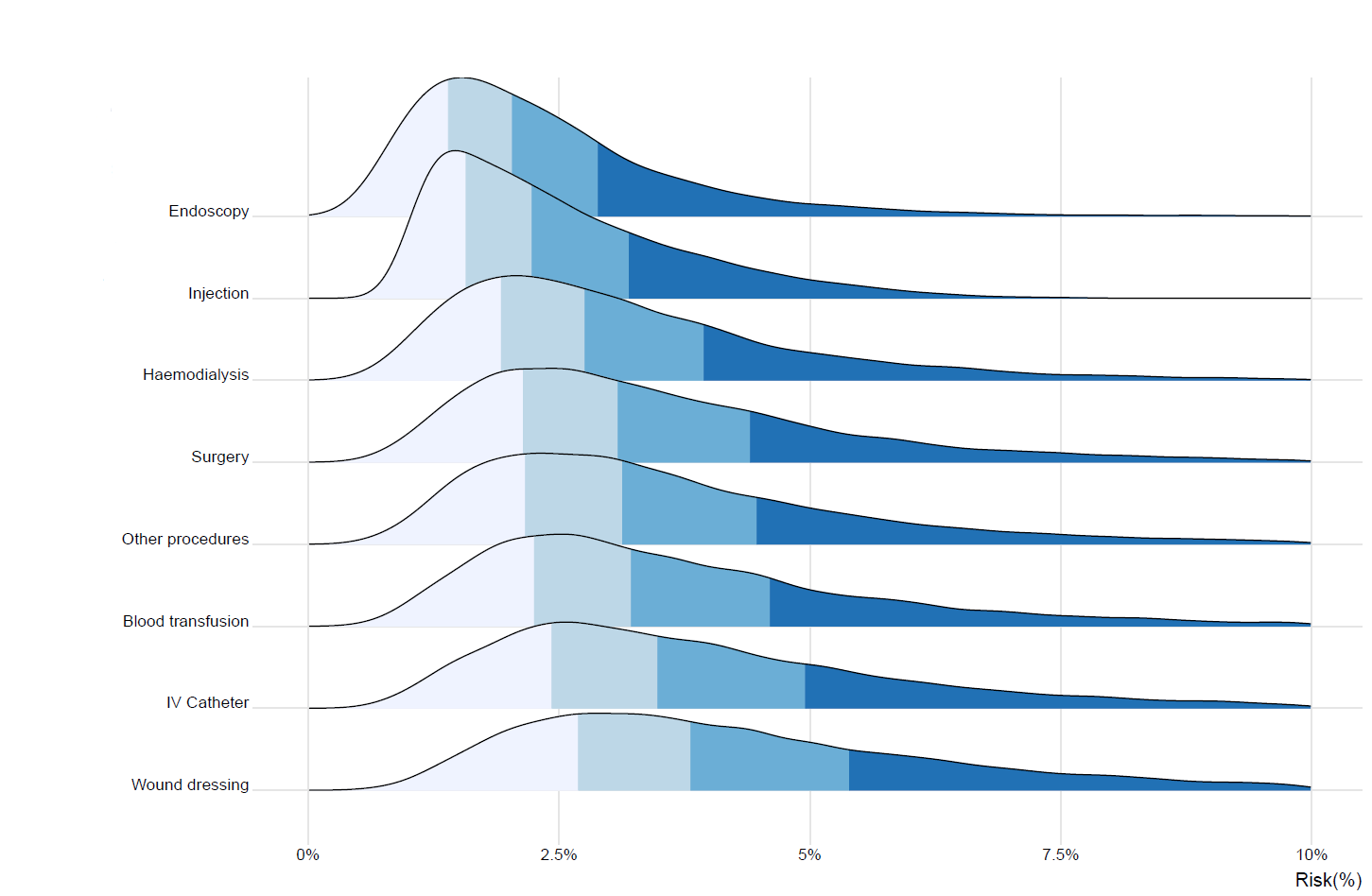

**Figure S4.** Distribution of the risk of HCV infection associated with different group of procedures considered in our work (Henriot *et al.*, 2022)

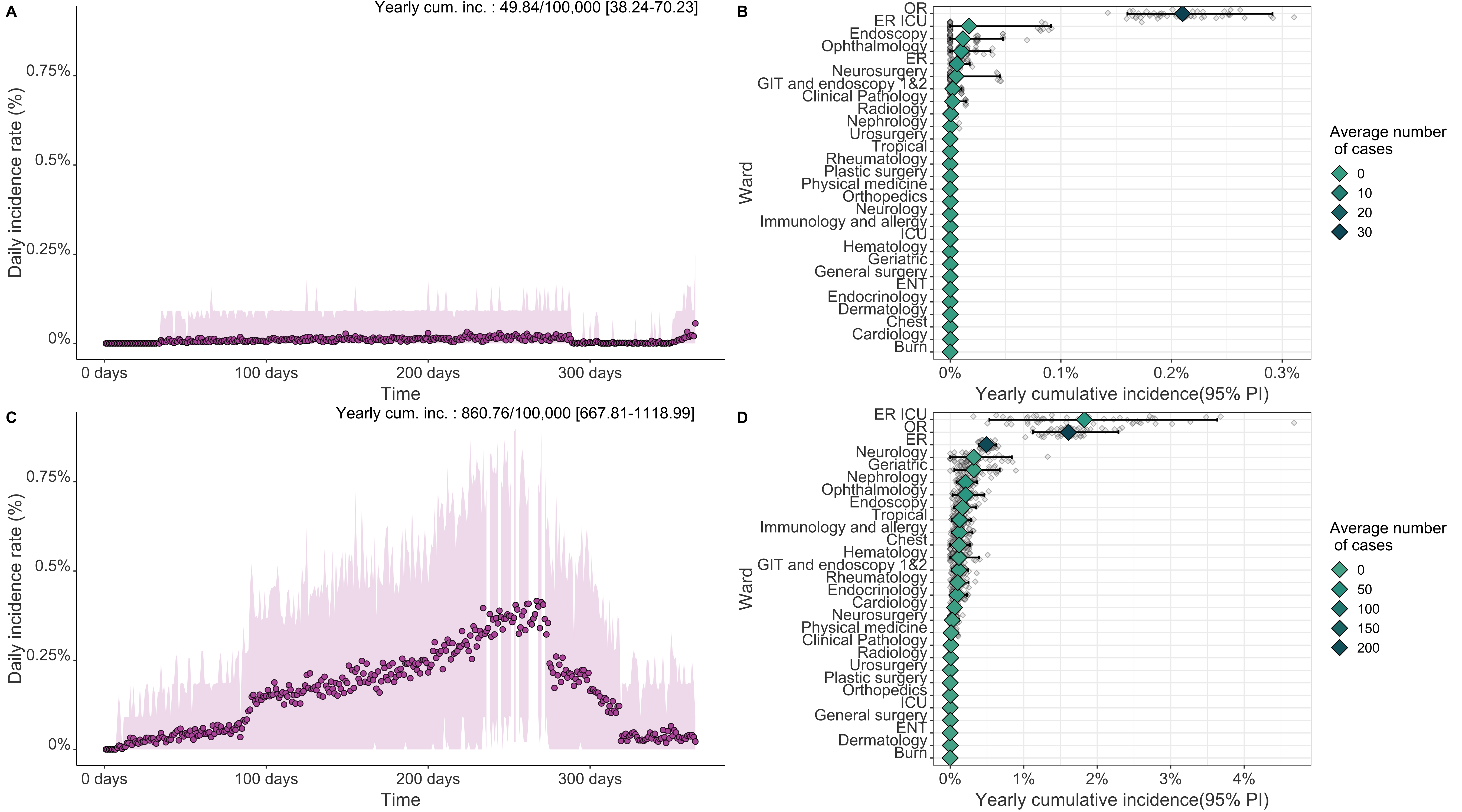

**Figure S5.** Results of the model for baseline scenarios (HBV case). (A) and (B): Daily incidence rate for the University hospital and private sector hospital, respectively. (C) and (D) Yearly cumulative incidence (mean and 95% PI) and average number of cases for each ward, ranked by mean cumulative incidence values.

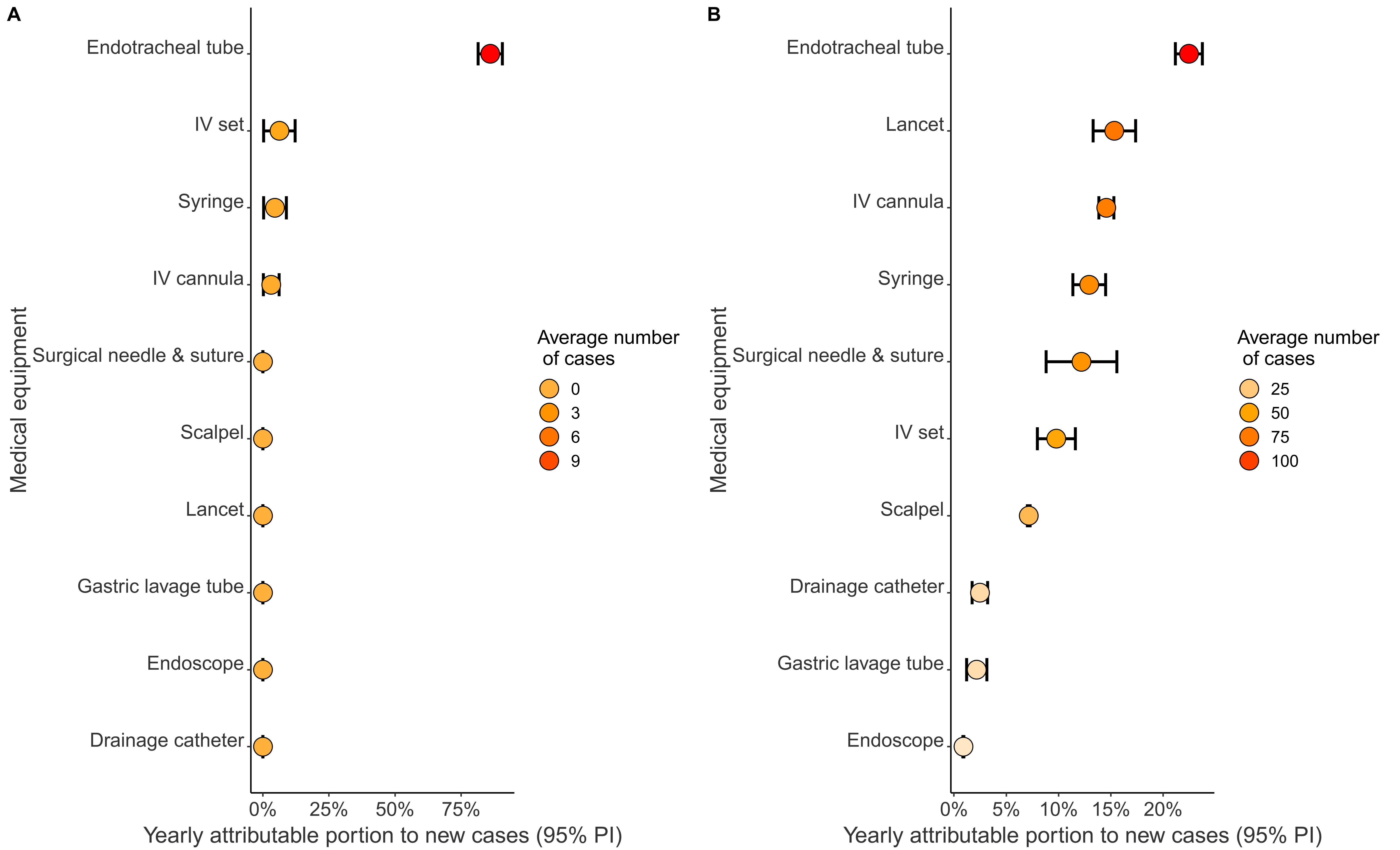

**Figure S6.** Yearly attributable portion to new cases for each device, in (A) Baseline scenario for the University Hospital and (B) Baseline scenario for the non-governmental hospital.

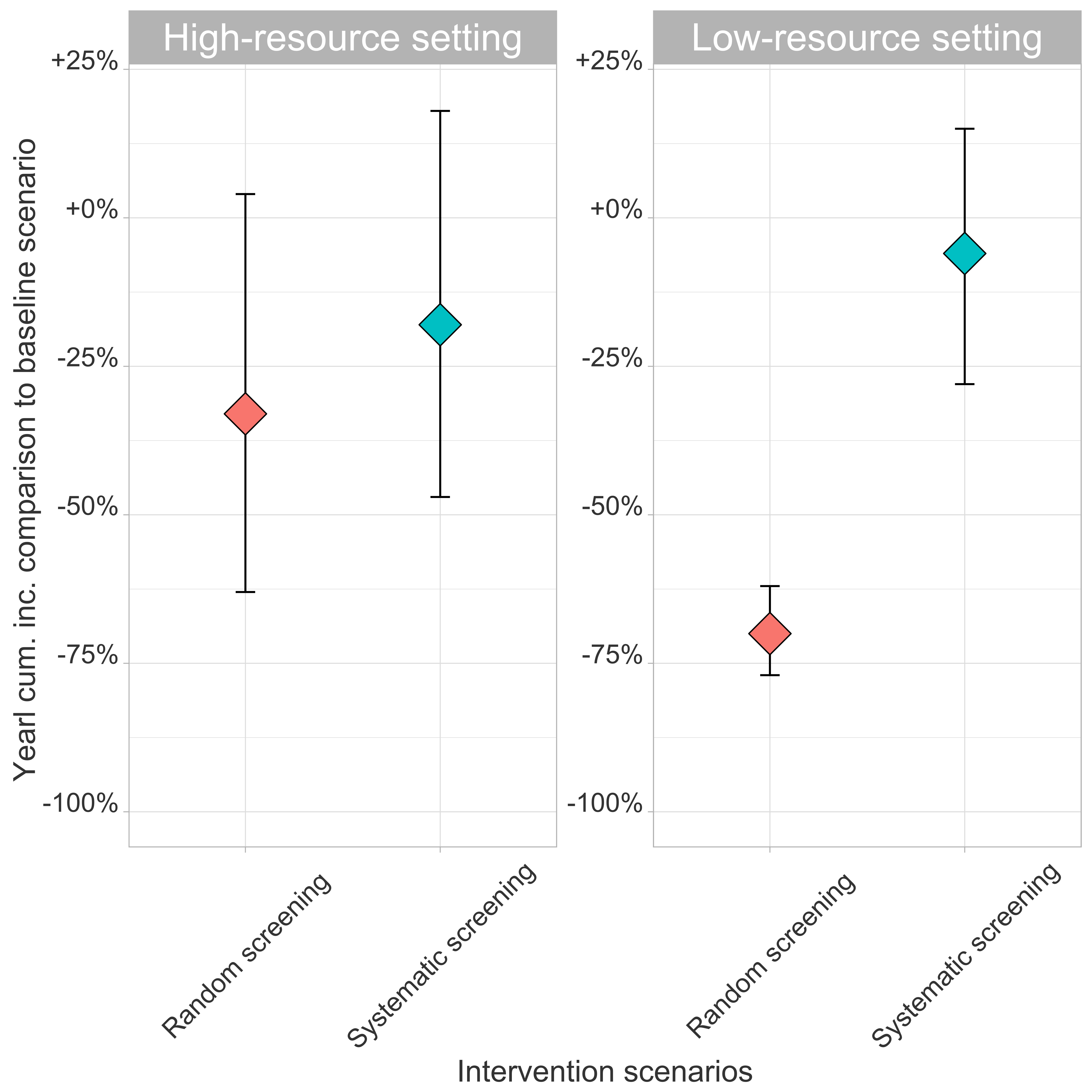

**Figure S7.** Yearly cumulative incidence for two different intervention strategies for (A) High-resource and (B) Low-resource hospitals. Baseline scenarios corresponds to the no-intervention scenarios. For the high-resource and low-resource settings on average of 30,380 patients (40.5%) and 53,472 (71%) patients were screened, respectively, (i) either systematically (in the three most at-risk wards) or (ii) randomly upon admission.

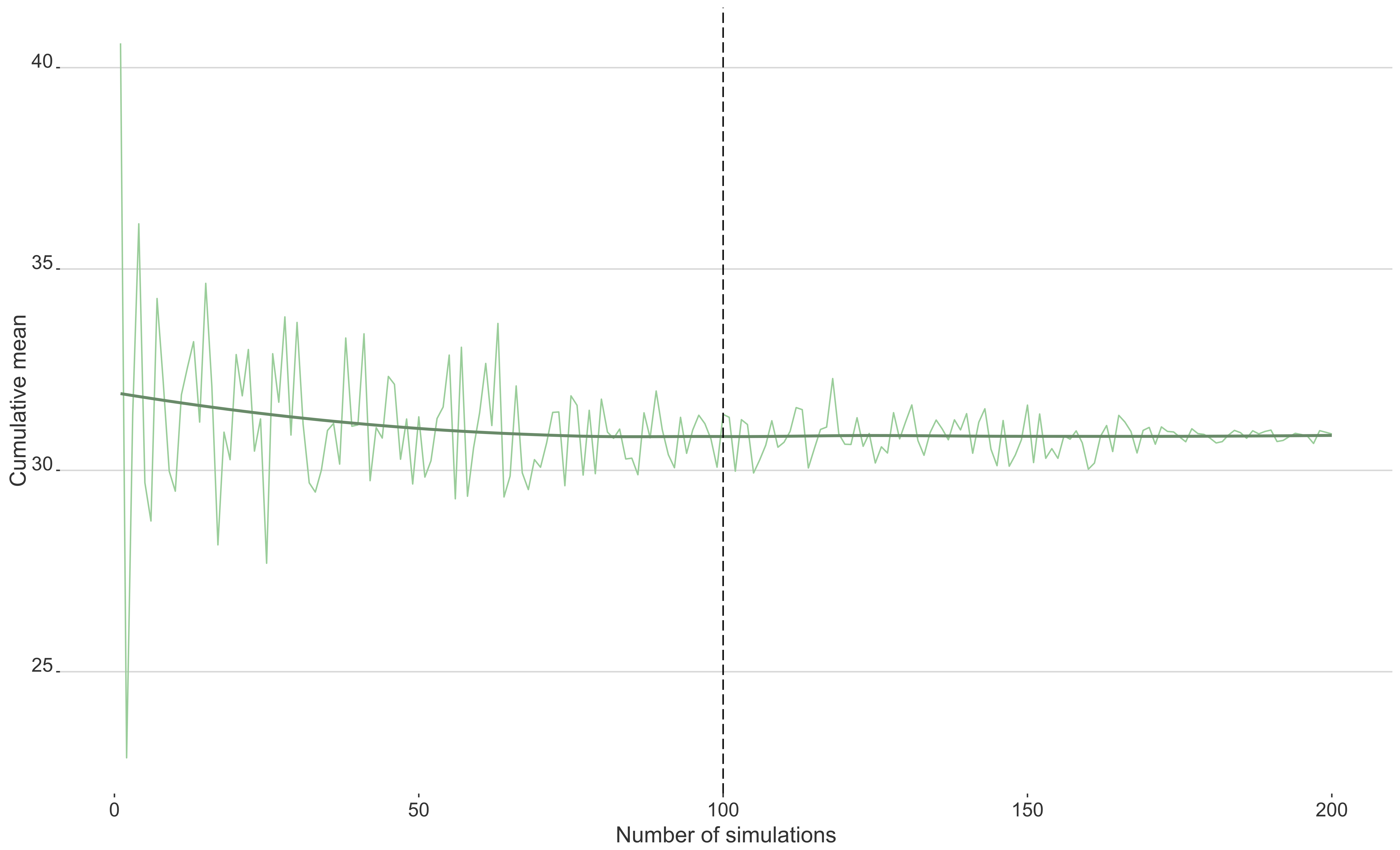

**Figure S8.** Cumulative mean of the number of annual cases for 1 to 200 simulations. This graph shows that the mean seems to converge after 100 simulations.
